# Overlap of high-risk individuals across family history, genetic & non-genetic breast cancer risk models: Analysis of 180,398 women from European & Asian ancestries

**DOI:** 10.1101/2025.02.27.25323002

**Authors:** Peh Joo Ho, Christine Kim Yan Loo, Meng Huang Goh, Mustapha Abubakar, Thomas U. Ahearn, Irene L. Andrulis, Natalia N. Antonenkova, Kristan J. Aronson, Annelie Augustinsson, Sabine Behrens, Clara Bodelon, Natalia V. Bogdanova, Manjeet K. Bolla, Kristen Brantley, Hermann Brenner, Helen Byers, Nicola J. Camp, Jose E. Castelao, Melissa H. Cessna, Jenny Chang-Claude, Stephen J. Chanock, Georgia Chenevix-Trench, Ji-Yeob Choi, Sarah V. Colonna, Kamila Czene, Mary B. Daly, Francoise Derouane, Thilo Dörk, A. Heather Eliassen, Christoph Engel, Mikael Eriksson, D. Gareth Evans, Olivia Fletcher, Lin Fritschi, Manuela Gago-Dominguez, Jeanine M. Genkinger, Willemina R.R. Geurts-Giele, Gord Glendon, Per Hall, Ute Hamann, Cecilia Y.S. Ho, Weang-Kee Ho, Maartje J. Hooning, Reiner Hoppe, Anthony Howell, Keith Humphreys, ABCTB Investigators, kConFab Investigators, SGBCC Investigators, MyBrCa Investigators, Hidemi Ito, Motoki Iwasaki, Anna Jakubowska, Helena Jernström, Esther M. John, Nichola Johnson, Daehee Kang, Sung-Won Kim, Cari M. Kitahara, Yon-Dschun Ko, Peter Kraft, Ava Kwong, Diether Lambrechts, Susanna Larsson, Shuai Li, Annika Lindblom, Martha Linet, Jolanta Lissowska, Artitaya Lophatananon, Robert J. MacInnis, Arto Mannermaa, Siranoush Manoukian, Sara Margolin, Keitaro Matsuo, Kyriaki Michailidou, Roger L. Milne, Nur Aishah Mohd Taib, Kenneth Muir, Rachel A. Murphy, William G. Newman, Katie M. O’Brien, Nadia Obi, Olufunmilayo I. Olopade, Mihalis I. Panayiotidis, Sue K. Park, Tjoung-Won Park-Simon, Alpa V. Patel, Paolo Peterlongo, Dijana Plaseska-Karanfilska, Katri Pylkäs, Muhammad U. Rashid, Gad Rennert, Juan Rodriguez, Emmanouil Saloustros, Dale P. Sandler, Elinor J. Sawyer, Christopher G. Scott, Shamim Shahi, Xiao-Ou Shu, Katerina Shulman, Jacques Simard, Melissa C. Southey, Jennifer Stone, Jack A. Taylor, Soo-Hwang Teo, Lauren R. Teras, Mary Beth Terry, Diana Torres, Celine M. Vachon, Maxime Van Houdt, Jelle Verhoeven, Clarice R. Weinberg, Alicja Wolk, Taiki Yamaji, Cheng Har Yip, Wei Zheng, Mikael Hartman, Jingmei Li

**Affiliations:** Human Genetics Division, Genome Institute of Singapore, Agency for Science, Technology and Research (A*STAR), Singapore, Singapore City, 138672; Saw Swee Hock School of Public Health, National University of Singapore, Singapore, Singapore City, 117549; Division of Cancer Epidemiology and Genetics, National Cancer Institute, National Institutes of Health, Department of Health and Human Services, USA, Bethesda, MD, 20850; Fred A. Litwin Center for Cancer Genetics, Lunenfeld-Tanenbaum Research Institute of Mount Sinai Hospital, Canada, Toronto, Ontario, M5G 1X5; Department of Molecular Genetics, University of Toronto, Canada, Toronto, Ontario, M5S 1A8; N.N. Alexandrov Research Institute of Oncology and Medical Radiology, Belarus, Minsk, 223040; Department of Public Health Sciences, and Sinclair Cancer Research Institute, Queen’s University, Canada, Kingston, ON, K7L 3N6; Oncology, Department of Clinical Sciences in Lund, Lund University, Sweden, Lund, 221 85; Division of Cancer Epidemiology, German Cancer Research Center (DKFZ), Germany, Heidelberg, 69120; Department of Population Science, American Cancer Society, USA, Atlanta, GA, 30303; Department of Radiation Oncology, Hannover Medical School, Germany, Hannover, 30625; Gynaecology Research Unit, Hannover Medical School, Germany, Hannover, 30625; Centre for Cancer Genetic Epidemiology, Department of Public Health and Primary Care, University of Cambridge, UK, Cambridge, CB1 8RN; Department of Epidemiology, Harvard T.H. Chan School of Public Health, USA, Boston, MA, 02115; Division of Clinical Epidemiology and Aging Research, German Cancer Research Center (DKFZ), Germany, Heidelberg, 69120; German Cancer Consortium (DKTK), German Cancer Research Center (DKFZ), Germany, Heidelberg, 69120; Division of Evolution and Genomic Sciences, School of Biological Sciences, Faculty of Biology, Medicine and Health, University of Manchester, Manchester Academic Health Science Centre, UK, Manchester, M13 9WL; Department of Internal Medicine and Huntsman Cancer Institute, University of Utah, USA, Salt Lake City, UT, 84112; Oncology and Genetics Unit, Instituto de Investigación Sanitaria de Santiago de Compostela (IDIS) Foundation, Complejo Hospitalario Universitario de Santiago, SERGAS, Spain, Vigo, 36312; Intermountain Healthcare, USA, Salt Lake City, UT, 84143; Cancer Epidemiology Group, University Cancer Center Hamburg (UCCH), University Medical Center Hamburg-Eppendorf, Germany, Hamburg, 20246; Cancer Research Program, QIMR Berghofer Medical Research Institute Vol. Locked Bag 2000, Herston, QLD 4029, Australia, Brisbane, Queensland, 4006; Department of Biomedical Sciences, Seoul National University Graduate School, Korea, Seoul, 03080; Cancer Research Institute, Seoul National University, Korea, Seoul, 03080; Institute of Health Policy and Management, Seoul National University Medical Research Center, Korea, Seoul, 03080; Department of Medical Epidemiology and Biostatistics, Karolinska Institutet, Sweden, Stockholm, 171 65; Department of Clinical Genetics, Fox Chase Cancer Center, USA, Philadelphia, PA, 19111; Leuven Multidisciplinary Breast Center, Department of Oncology, Leuven Cancer Institute, University Hospitals Leuven, Belgium, Leuven, 3000; Channing Division of Network Medicine, Department of Medicine, Brigham and Women’s Hospital and Harvard Medical School, USA, Boston, MA, 02115; Department of Nutrition, Harvard T.H. Chan School of Public Health, USA, Boston, MA, 02115; Institute for Medical Informatics, Statistics and Epidemiology, University of Leipzig, Germany, Leipzig, 04107; LIFE - Leipzig Research Centre for Civilization Diseases, University of Leipzig, Germany, Leipzig, 04103; North West Genomics Laboratory Hub, Manchester Centre for Genomic Medicine, St Mary’s Hospital, Manchester University NHS Foundation Trust, Manchester Academic Health Science Centre, UK, Manchester, M13 9WL; The Breast Cancer Now Toby Robins Research Centre, The Institute of Cancer Research, UK, London, SW7 3RP; School of Population Health, Curtin University, Australia, Perth, Western Australia, 6102; Cancer Genetics and Epidemiology Group, Genomic Medicine Group, Fundación Instituto de Investigación Sanitaria de Santiago de Compostela (FIDIS), Complejo Hospitalario Universitario de Santiago, SERGAS, Spain, Santiago de Compostela, 15706; Department of Epidemiology, Mailman School of Public Health, Columbia University, USA, New York, NY, 10032; Herbert Irving Comprehensive Cancer Center, USA, New York, NY; Department of Clinical Genetics, Erasmus University Medical Center Vol. P.O. Box 2040, 3000 CA, the Netherlands, Rotterdam, 3015 CN; Department of Oncology, Södersjukhuset, Sweden, Stockholm, 118 83; Molecular Genetics of Breast Cancer, German Cancer Research Center (DKFZ), Germany, Heidelberg, 69120; Department of Molecular Pathology, Hong Kong Sanatorium and Hospital, Hong Kong; School of Mathematical Sciences, Faculty of Science and Engineering, University of Nottingham Malaysia, Malaysia, Semenyih, Selangor, 43500; Cancer Research Malaysia, Malaysia, Subang Jaya, Selangor, 47500; Department of Medical Oncology, Erasmus MC Cancer Institute Vol. P.O. Box 2040, 3000 CA, the Netherlands, Rotterdam, 3015 GD; Dr. Margarete Fischer-Bosch-Institute of Clinical Pharmacology, Germany, Stuttgart, 70376; University of Tübingen, Germany, Tübingen, 72074; Division of Cancer Sciences, University of Manchester, UK, Manchester, M13 9PL; Australian Breast Cancer Tissue Bank, Westmead Institute for Medical Research, University of Sydney, Australia, Sydney, New South Wales, 2145; Research Department, Peter MacCallum Cancer Center, Australia, Melbourne, Victoria, 3000; Sir Peter MacCallum Department of Oncology, The University of Melbourne, Australia, Parkville, Victoria, 3010; Department of Surgery, National University Health System, Singapore, Singapore City, 119228; Department of Surgery, Yong Loo Lin School of Medicine, National University of Singapore, Singapore, Singapore City, 119077; Cancer Genetics Service, National Cancer Centre, Singapore, Singapore City, 169610; Breast Department, KK Women’s and Children’s Hospital, Singapore, Singapore City, 229899; SingHealth Duke-NUS Breast Centre, Singapore, Singapore City, 168753; Department of General Surgery, Tan Tock Seng Hospital, Singapore, Singapore City, 308433; Division of Surgery and Surgical Oncology, National Cancer Centre, Singapore, Singapore City, 169610; Department of General Surgery, Singapore General Hospital, Singapore, Singapore City, 169608; Division of Breast Surgery, Department of General Surgery, Changi General Hospital, Singapore, Singapore City, 529889; Division of Radiation Oncology, National Cancer Centre, Singapore, Singapore City, 169610; Division of Medical Oncology, National Cancer Centre, Singapore, Singapore City, 169610; Breast Cancer Research Unit, University Malaya Cancer Research Institute, Faculty of Medicine, University of Malaya, Malaysia, Kuala Lumpur, 50603; Division of Cancer Information and Control, Aichi Cancer Center Research Institute, Japan, Nagoya, 464-8681; Division of Cancer Epidemiology, Nagoya University Graduate School of Medicine, Japan, Nagoya, 466-8550; Division of Epidemiology, National Cancer Center Institute for Cancer Control, Japan, Tokyo, 104-0045; Independent Laboratory of Molecular Biology and Genetic Diagnostics, Pomeranian Medical University, Poland, Szczecin, 171-252; International Hereditary Cancer Center, Department of Genetics and Pathology, Pomeranian Medical University in Szczecin, Poland, Szczecin, 70-115; Department of Epidemiology and Population Health, Stanford University School of Medicine, USA, Stanford, CA, 94305; Department of Medicine, Division of Oncology, Stanford Cancer Institute, Stanford University School of Medicine, USA, Stanford, CA, 94304; Department of Preventive Medicine, Seoul National University College of Medicine, Korea, Seoul, 03080; Department of Surgery, Daerim Saint Mary’s Hospital, Korea, Seoul, 07442; Radiation Epidemiology Branch, Division of Cancer Epidemiology and Genetics, National Cancer Institute, USA, Bethesda, MD, 20892; Department of Internal Medicine, Johanniter GmbH Bonn, Johanniter Krankenhaus, Germany, Bonn, 53177; Hong Kong Hereditary Breast Cancer Family Registry, Hong Kong; Department of Surgery, The University of Hong Kong, Hong Kong; Department of Surgery and Cancer Genetics Center, Hong Kong Sanatorium and Hospital, Hong Kong; Laboratory for Translational Genetics, Department of Human Genetics, KU Leuven, Belgium, Leuven, 3000; VIB Center for Cancer Biology, VIB, Belgium, Leuven, 3001; Institute of Environmental Medicine, Karolinska Institutet, Sweden, Stockholm, 171 77; Department of Surgical Sciences, Uppsala University, Sweden, Uppsala, 751 05; Centre for Epidemiology and Biostatistics, Melbourne School of Population and Global Health, The University of Melbourne, Australia, Melbourne, Victoria, 3010; Precision Medicine, School of Clinical Sciences at Monash Health, Monash University, Australia, Clayton, Victoria, 3168; Department of Molecular Medicine and Surgery, Karolinska Institutet, Sweden, Stockholm, 171 76; Department of Clinical Genetics and Genomics, Karolinska University Hospital, Sweden, Stockholm, 171 76; Department of Cancer Epidemiology and Prevention, M. Sklodowska-Curie National Research Oncology Institute, Poland, Warsaw, 02-034; Division of Population Health, Health Services Research and Primary Care, School of Health Sciences, Faculty of Biology, Medicine and Health, The University of Manchester, UK, Manchester, M13 9PL; Cancer Epidemiology Division, Cancer Council Victoria, Australia, Melbourne, Victoria, 3004; Translational Cancer Research Area, University of Eastern Finland, Finland, Kuopio, 70210; Institute of Clinical Medicine, Pathology and Forensic Medicine, University of Eastern Finland, Finland, Kuopio, 70210; Biobank of Eastern Finland, Kuopio University Hospital, Finland, Kuopio; Unit of Medical Genetics, Department of Medical Oncology and Hematology, Fondazione IRCCS Istituto Nazionale dei Tumori di Milano, Italy, Milan, 20133; Department of Clinical Science and Education, Södersjukhuset, Karolinska Institutet, Sweden, Stockholm, 118 83; Division of Cancer Epidemiology and Prevention, Aichi Cancer Center Research Institute, Japan, Nagoya, 464-8681; Biostatistics Unit, The Cyprus Institute of Neurology and Genetics Vol. P.O.Box 23462, 1683, Nicosia, Cyprus, Cyprus, Nicosia, 2371; Department of Surgery, Faculty of Medicine, University of Malaya, UM Cancer Research Institute, Malaysia, Kuala Lumpur, 50603; School of Population and Public Health, University of British Columbia, Canada, Vancouver, BC, V6T 1Z4; Cancer Control Research, BC Cancer Agency, Canada, Vancouver, BC, V5Z 1L3; Epidemiology Branch, National Institute of Environmental Health Sciences, NIH, USA, Research Triangle Park, NC, 27709; Institute for Occupational and Maritime Medicine, University Medical Center Hamburg-Eppendorf, Germany, Hamburg, 20246; Institute for Medical Biometry and Epidemiology, University Medical Center Hamburg-Eppendorf, Germany, Hamburg, 20246; Center for Clinical Cancer Genetics, The University of Chicago, USA, Chicago, IL, 60637; Department of Cancer Genetics, Therapeutics and Ultrastructural Pathology, The Cyprus Institute of Neurology & Genetics Vol. P.O.Box 23462, 1683, Nicosia, Cyprus, Cyprus, Nicosia, 2371; Integrated Major in Innovative Medical Science, Seoul National University College of Medicine, Korea, Seoul, 03080; Genome Diagnostics Program, IFOM ETS - the AIRC Institute of Molecular Oncology, Italy, Milan, 20139; Research Centre for Genetic Engineering and Biotechnology ‘Georgi D. Efremov’, MASA, Republic of North Macedonia, Skopje, 1000; Laboratory of Cancer Genetics and Tumor Biology, Translational Medicine Research Unit, Biocenter Oulu, University of Oulu, Finland, Oulu, 90220; Laboratory of Cancer Genetics and Tumor Biology, Northern Finland Laboratory Centre Oulu, Finland, Oulu, 90220; Department of Basic Sciences, Shaukat Khanum Memorial Cancer Hospital and Research Centre (SKMCH & RC), Pakistan, Lahore, 54000; Faculty of Medicine, Technion – Israel Institute of Technology and Association for Promotion of Research in Precision Medicine, Israel, Haifa, 35254; Division of Oncology, Faculty of Medicine, School of Health Sciences, University of Thessaly, Greece, Larissa, 411 10; School of Cancer & Pharmaceutical Sciences, Comprehensive Cancer Centre, Guy’s Campus, King’s College London, UK, London; Department of Quantitative Health Sciences, Division of Clinical Trials and Biostatistics, Mayo Clinic, USA, Rochester, MN, 55905; Division of Epidemiology, Department of Medicine, Vanderbilt Epidemiology Center, Vanderbilt-Ingram Cancer Center, Vanderbilt University School of Medicine, USA, Nashville, TN, 37232; Clalit Regional Oncology Unit, Haifa and Western Galilee District, Israel, Haifa; Genomics Center, Centre Hospitalier Universitaire de Québec – Université Laval Research Center, Canada, Québec City, Québec, G1V 4G2; Department of Clinical Pathology, The University of Melbourne, Australia, Melbourne, Victoria, 3010; Genetic Epidemiology Group, School of Population and Global Health, University of Western Australia, Australia, Perth, Western Australia, 6009; Epigenetic and Stem Cell Biology Laboratory, National Institute of Environmental Health Sciences, NIH, USA, Research Triangle Park, NC, 27709; Institute of Human Genetics, Pontificia Universidad Javeriana, Colombia, Bogota, 110231; Department of Quantitative Health Sciences, Division of Epidemiology, Mayo Clinic, USA, Rochester, MN, 55905; Biostatistics and Computational Biology Branch, National Institute of Environmental Health Sciences, NIH, USA, Research Triangle Park, NC, 27709; Subang Jaya Medical Centre, Malaysia, Subang Jaya, Selangor, 47500; National Cancer Centre, Singapore, Singapore City, 168583

**Keywords:** Breast cancer, Ductal Carcinoma In Situ (DCIS), Polygenic risk score (PRS), Gail model, Risk stratification, BRCA1, BRCA2, risk-based screening

## Abstract

**Background:** Breast cancer is multifactorial. Focusing on limited risk factors may miss high-risk individuals.

**Methods:** We assessed the performance and overlap of various risk factors in identifying high-risk individuals for invasive breast cancer (BrCa) and ductal carcinoma in situ (DCIS) in 161,849 European-ancestry and 18,549 Asian-ancestry women. Discriminatory ability was evaluated using the area under the receiver operating characteristic curve (AUC). High-risk criteria included: 5-year absolute risk ≥1·66% by the Gail model [GAIL_binary_]; first-degree family history of breast cancer [FH_binary_]; 5-year absolute risk ≥1·66% by a 313-variants polygenic risk score [PRS_binary_]; and carriers of pathogenic variants in breast cancer predisposition genes [PTV_binary_].

**Findings:** The 5-year absolute risk by PRS outperformed the Gail model in predicting BrCa (Europeans_vs controls_: AUC_PRS_=0·635 [0·632-0·638] vs AUC_Gail_=0·492 [0·489-0·495]; Asians_vs controls_: AUC_PRS_=0·564 [0·556-0·573] vs AUC_Gail_=0·506 [0·497-0·514]). PRS_binary_ and GAIL_binary_ identified more high-risk European than Asia individuals. High-risk proportions were higher among BrCa (16-26%) and DCIS (20-33%) compared to controls (9-15%) among young Europeans and all Asians. Fewer than 7% of BrCa, 10% of DCIS, and 3% of controls were classified as high-risk by multiple risk classifiers. Overlap between PRS_binary_ and PTV_binary_ was minimal (<0·65% Europeans, <0·15% Asians) compared to the proportion at high risk using PTV_binary_ alone (Europeans: 4·6%, Asians: 4·4%) and PRS_binary_ alone (Europeans: 13·9%, Asians: 8·5%). PRS_binary_ and FH_binary_ uniquely identified 5-6% and 9-11% of young BrCa, respectively.

**Interpretation:** The incomplete overlap between high-risk individuals identified by PRS_binary_, GAIL_binary_, FH_binary,_ and PTV_binary_ highlights the need for a comprehensive approach to breast cancer risk prediction.

**SIGNIFICANCE:** This study shows that different ways of predicting breast cancer risk do not always flag the same people, suggesting that combining multiple risk factors could improve early detection and screening.

## INTRODUCTION

A worldwide increase of 31% in the number of breast cancer cases is projected over the next two decades.^1^ Early detection significantly improves survival rates.^2,3^ Multiple studies have shown that mammogram screenings reduce mortality rates for women above 50 years of age, while the benefits of screening for those younger are less clear.^4,5^ Current screening guidelines are based on age, yet many patients are diagnosed before reaching the recommended screening age.^6^

Advances in breast cancer research suggest the potential for more risk-based approaches to cost-effective screening programs.^7^ Developed in the 1980s, the Gail model, a validated statistical tool, uses personal information to estimate breast cancer risk over the next five years. Originally developed for White females in the United States without a history of in situ or invasive breast cancer, its accuracy for non-White populations is debated.^8–10^ Common issues include both underestimation and overestimation in non-European populations, leading to unclear recommendations for diverse ethnic groups.^9,11^

In addition to non-genetic risk factors, studies have explored the use of polygenic risk scores (PRS) to enhance existing prediction models.^12,13^ Breast cancer has a significant heritable component. While PRS has added value to prediction models, its implementation, particularly in Asian populations, remains inconclusive.^15^ This is partly because PRS training datasets have predominantly included European populations due to their larger representation in research.^16^

Protein-truncating variants (PTVs) are another genetic factor used in risk prediction. Unlike PRS, which aggregates the associated effects of numerous, relatively common genetic variants across the genome, PTVs specifically target variants that lead to premature protein termination, potentially disrupting gene function. This distinction means that PTVs are specific genetic changes with known functional impacts, while PRS provides an overview of associated genetic risks of small effect sizes across the genome. PTVs in nine breast cancer predisposition genes, *ATM, BRCA1, BRCA2, CHEK2, PALB2, BARD1, RAD51C, RAD51D,* or *TP53* have been shown clinically useful for inclusion on breast cancer risk prediction panels in a large analytical cohort comprising over 113,000 subjects and has been included in BOADICEA (Breast and Ovarian Analysis of Disease Incidence and Carrier Estimation Algorithm).^17,18^

Studying the overlap of genetic and non-genetic risk factors in identifying high-risk individuals will provide us with information on complementary risk factors that will enhance our ability to identify the subgroup of the population who would benefit from risk reduction interventions. In this case-control analysis involving the Breast Cancer Consortium (BCAC) dataset, we explore how prediction tools— such as the Gail model, PRS, PTVs in known breast cancer predisposition genes, and family history— apply to both European and Asian populations across non-screening and screening age groups.

## METHODS

### Study population

BCAC is an international collaboration that was formed to provide large sample sizes for investigating genetic associations.^19^ Women diagnosed with invasive breast cancer (BrCa) or ductal carcinoma in situ patients (DCIS), and women with no prior diagnosis of breast cancer (controls) were recruited by study groups across the globe and collectively studied under BCAC.^20^ Our study focuses on individuals who are genetically Asian or European-White (from here on referred to as “European”).

To reduce the influence of missing values on the performance of the Gail model, studies with missing values for 50% or more for each of at least two of the three risk factors in the Gail model^8^ –age of menarche, age at first live birth, and first-degree family history of breast cancer–were excluded. The studies included are listed in **Supplementary Table 1·** Exclusion was done separately for individual studies and each disease status (BrCa, DCIS, and controls).

Further exclusions were made on an individual level (**Supplementary Figure 1**). Women with unknown age at enrolment for controls (n=5,566) and unknown age at diagnosis for BrCa and DCIS cases (n=2,103) were excluded. Women below the age of 30 years (n=2,360) and women above 80 years (n=1,897) for whom the Gail model prediction is not valid were excluded. A total of 180,398 individuals were included in our study. We compared demographic differences between the included and excluded individuals to assess potential selection bias. The result is presented in **Additional Material - Supplementary Table 2·**

### Criteria to identify women at high risk of breast cancer

Four criteria were used to identify women at high risk of breast cancer: 1) 5-year absolute risk ≥1·66% by the Gail model [GAIL_binary_], 2) first-degree family history for breast cancer [FH_binary_, yes/no], 3) 5-year absolute risk ≥1·66% by a 313-variant breast cancer polygenic risk score^14^ [PRS_binary_], and in a subset of women 4) carriers of pathogenic variants in breast cancer predisposition genes [PTV_binary_]. The 1.66% five-year absolute risk threshold for breast cancer is widely adopted in clinical and research settings to reflect the level of risk at which women are considered for additional screening or preventive interventions, such as tamoxifen or raloxifene (from here onwards the high-risk category).^21^ Details of each risk factor are presented in **Additional Materials - Methods ·**

Due to the large number of studies with varying degrees of missing data for different risk factors, the parsimonious Gail model, which most studies would have information on, was selected.^8^ The R package “BCRA” (version 2.1.2) was used to calculate 5-year absolute risk.^8^ Implementation is described in Additional Material-Genetic breast cancer risk factor. In our analysis, those with unknown family history were considered to have no family history.

We studied genetic risk based on common germline variants associated with breast cancer, using the breast cancer PRS with 313 variants calculated with PLINK (version 3) with the scoresum option.^22,23^ The 5-year absolute risk was calculated by estimating the theoretical odds ratio of this percentile in relation to the 40-60 percentile, which is taken to represent the general population.^24^ A subgroup of individuals (n_European_=56,387, n_Asian_=3,617) had both genotyping and targeted-sequencing data. Nine breast cancer predisposition genes (PTVs in *ATM, BRCA1, BRCA2, CHEK2, PALB2, BARD1, RAD51C, RAD51D,* or *TP53*) were studied collectively.

#### Statistical analysis

Differences in genetic and non-genetic breast cancer risk factors of BrCa, DCIS, and controls were assessed using the Chi-squared test (categorical variables) and Kruskal-Wallis test (continuous variables). Venn diagrams (R package “VennDiagram”) were used to visualise the overlaps in high-risk individuals identified by the high-risk criteria. We subset the population by ancestry (European or Asian) and age (30-49 or 50-80 years). Overlaps between pairs of high-risk criteria were further considered by country. In the subset of individuals with both genotyping and targeted sequencing information (n=60,004), PRS_binary_ and PTV_binary_ (yes/no) were analysed for their ability to uniquely identify high-risk individuals.

### Evaluating the drivers of Gail risk score in differentiating breast cancer cases from controls

Although studies with high missingness rates for variables required to compute the Gail risk score were excluded (see “Excluded participants” above), there were still individuals with missing values. Hence, we studied the potential drivers of the Gail risk score in discriminating BrCa cases from controls using logistic regression models. All combinations of risk factors, where available, were assessed. Discriminatory ability was assessed by the area under the receiver operator curve (AUC). Missing values were coded in accordance to the “BCRA”.^8^

All analyses were performed in R version 4.2.2.

## RESULTS

### Cohort description

A total of 180,398 women were included, where 161,849 (90%) women were of European-ancestry and 18,549 (10%) were Asian-ancestry (**Table 1**). Of the European women, 68,540 (42%) were controls, and 83,685 (52%) were diagnosed with BrCa. Of the 18,549 Asian women, 8,347 (45%) were controls and 9,222 (50%) were BrCa cases (**Table 1**). In addition, there were 9,624 (6%) and 980 (5%) DCIS cases in European and Asian women, respectively (**Table 1).**

**Table 1.** Characteristics of 161,849 European-ancestry and 18,549 Asian-ancestry individuals (non-breast cancer controls (controls), patients diagnosed with invasive breast cancer (BrCa) and patients diagnosed with DCIS (DCIS)) between ages 30 and 80 years. IQR interquartile range. Family history: number of first-degree relatives with breast cancer.

|  | European-ancestry, n=161,849 |  |  | Asian-ancestry, n=18,549 |  |  |
| --- | --- | --- | --- | --- | --- | --- |
|  | Controls,<br>n=68,540 (42%) | BrCa,<br>n=83,685 (52%) | DCIS,<br>n=9,624 (6%) | Controls,<br>n= 8,347 (45%) | BrCa,<br>n=9,222 (50%) | DCIS,<br>n=980 (5%) |
| **Median age (at interview/ diagnosis), years (IQR)** | 57 (50 to 64) | 57 (49 to 65) | 55 (50 to 63) | 50 (44 to 58) | 49 (43 to 57) | 49 (43 to 56) |
| **Age at menarche, n (%)** |  |  |  |  |  |  |
| <12 years | 9,825 (14) | 10,859 (13) | 1,733 (18) | 508 (6) | 586 (6) | 61 (6) |
| 12 to 14 years | 30,243 (44) | 33,476 (40) | 4,546 (47) | 3,080 (37) | 3,278 (36) | 326 (33) |
| >=14 years | 21,133 (31) | 23,169 (28) | 2,758 (29) | 4,321 (52) | 4,186 (45) | 467 (48) |
| Unknown | 7,339 (11) | 16,181 (19) | 587 (6) | 438 (5) | 1,172 (13) | 126 (13) |
| **Age at first full-term pregnancy, n (%)** |  |  |  |  |  |  |
| Nulliparous | 8,319 (12) | 10,769 (13) | 1,520 (16) | 1,005 (12) | 1,236 (13) | 158 (16) |
| <20 years | 5,427 (8) | 6,138 (7) | 794 (8) | 316 (4) | 362 (4) | 29 (3) |
| 20 to 24 years | 21,863 (32) | 21,927 (26) | 2,899 (30) | 1,948 (23) | 1,810 (20) | 135 (14) |
| 25 to 29 years | 17,543 (26) | 18,174 (22) | 2,382 (25) | 2,836 (34) | 3,030 (33) | 293 (30) |
| >=30 years | 8,487 (12) | 9,973 (12) | 1,386 (14) | 1,149 (14) | 1,554 (17) | 156 (16) |
| Unknown | 6,901 (10) | 16,704 (20) | 643 (7) | 1,093 (13) | 1,230 (13) | 209 (21) |
| **Family history, n (%)** |  |  |  |  |  |  |
| No | 45,629 (67) | 48,903 (58) | 4,005 (42) | 7,181 (86) | 7,500 (81) | 708 (72) |
| 1 | 5,348 (8) | 10,256 (12) | 1,115 (12) | 437 (5) | 876 (9) | 114 (12) |
| 2+ | 791 (1) | 2,064 (2) | 305 (3) | 63 (1) | 93 (1) | 13 (1) |
| Unknown | 16,772 (24) | 22,462 (27) | 4,199 (44) | 666 (8) | 753 (8) | 145 (15) |
| **Number of breast biopsy, n (%)** |  |  |  |  |  |  |
| No | 930 (1) | 3,181 (4) | 96 (1) | 0 (0) | 0 (0) | 0 (0) |
| 1 | 277 (0) | 3,148 (4) | 216 (2) | 0 (0) | 0 (0) | 0 (0) |
| 2+ | 103 (0) | 1,822 (2) | 148 (2) | 0 (0) | 0 (0) | 0 (0) |
| Unknown | 67,230 (98) | 75,534 (90) | 9,164 (95) | 8,347 (100) | 9,222 (100) | 980 (100) |
| **Atypical hyperplasia, n (%)** |  |  |  |  |  |  |
| No | 930 (1) | 3,181 (4) | 96 (1) | 0 (0) | 0 (0) | 0 (0) |
| Yes | 5 (0) | 49 (0) | 8 (0) | 0 (0) | 0 (0) | 0 (0) |
| Unknown | 67,605 (99) | 80,455 (96) | 9,520 (99) | 8,347 (100) | 9,222 (100) | 980 (100) |
| **Median 5-year absolute risk by Gail (IQR)** | 1.25 (0.92 to 1.64) | 1.25 (0.89 to 1.73) | 1.29 (0.98 to 1.71) | 0.61 (0.45 to 0.78) | 0.61 (0.44 to 0.80) | 0.61 (0.38 to 0.81) |
| **Protein truncating variants (9 Genes)** |  |  |  |  |  |  |
| No | 24,215 (35) | 27,926 (33) | 1,662 (17) | 1,091 (13) | 2,049 (22) | 317 (32) |
| Yes | 583 (1) | 1,927 (2) | 74 (1) | 24 (0) | 129 (1) | 7 (1) |
| Unknown | 43,742 (64) | 53,832 (64) | 7,888 (82) | 7,232 (87) | 7,044 (76) | 656 (67) |
| **Polygenic risk score (PRS)** | -0.45 (-0.86 to -0.04) | -0.09 (-0.51 to 0.32) | -0.15 (-0.55 to 0.27) | 0.16 (-0.20 to 0.53) | 0.37 (-0.01 to 0.76) | 0.45 (0.05 to 0.83) |
| **Median 5-year absolute risk by PRS (IQR)** | 0.69 (0.46 to 1.05) | 0.95 (0.62 to 1.44) | 0.91 (0.61 to 1.37) | 0.62 (0.40 to 0.95) | 0.74 (0.44 to 1.15) | 0.79 (0.44 to 1.28) |

### European-ancestry study population

The median age at diagnosis for BrCa cases of European ancestry was 57 years [interquartile range [IQR]: 49-65]. The corresponding age at interview for European-ancestry controls was 57 years [IQR: 50-64] (**Table 1**). BrCa cases were more likely to have a family history than controls (14% vs 9%, respectively). The distribution of the 5-year absolute risk by the Gail model was very similar between BrCa cases (median 1·25% [IQR: 0·89-1·73]) and controls (1·25% [IQR: 0·92-1·64]). The median 5-year absolute risk by PRS in BrCa cases was 0·95% [IQR: 0·62-1·44], significantly greater than that of controls 0·69% [IQR: 0·46-1·05]. The PRS distributions (scoresum) and 5-year absolute risks were similar across countries (**Supplementary Figure 2**). Less than 40% of the population had PTV information. Out of 29,853 European BrCa patients, 1,927 (6%) were PTV carriers, which is three times the proportion found in the control group (583 out of 24,798, or 2%). The observations were largely similar for DCIS (**Table 1**).

### Asian-ancestry study population

The median age at diagnosis for BrCa cases was 49 years [IQR: 43-57], and the age at enrolment was 50 years [IQR: 44-58] for controls (**Table 1**). Of the BrCa cases, 10% reported positive family history, while a smaller proportion of controls (6%) reported so. The distribution of 5-year absolute risk by the Gail model was not significantly different between BrCa cases and controls. The distribution of 5-year absolute risk by PRS for BrCa patients was shifted rightwards of controls (p<0.001). The distribution of PRS (scoresum) and 5-year absolute risks varied by country (**Supplementary Figure 3).** Among BrCa patients with known PTV information (n=2,178) 6% were mutation carriers. A smaller percentage (2%) of controls (n=1,115) were PTV carriers. As with the Europeans, the observations were mostly similar for DCIS in Asians **(Table 1)**. and controls.

### Associations between risk stratifiers, BrCa and DCIS

**Table 2** and **Table 3** display the strengths of association of different risk stratifiers (PRS_binary_, GAIL_binary_, and FH_binary_) with BrCa and DCIS, respectively, stratified by ancestry and age groups. Using both PRS_binary_ and GAIL_binary_ (i.e. individuals is stratified as high risk when either PRS or GAIL is ≥1.66%) improves the discriminatory ability as compared to using GAIL_binary_ alone (in Europeans: AUC_BrCa-PRS_GAIL_=0·554 [0·552 to 0·557] vs AUC_BrCa-GAIL_=0·522 [0·520-0·524]; in Asian: AUC_BrCa-PRS_GAIL_=0·527 [0·523 to 0·532] vs AUC_BrCa-GAIL_=0·506 [0·503-0·508]) (**Table 2**), In Europeans, the odds ratios and corresponding 95% confidence intervals for PRS_binary_ and GAIL_binary_ (≥1·66% 5-year absolute risk threshold) were 2·60 [2·52-2·69] and 1·25 [1·22-1·28], respectively, for BrCa, and 2·21 [2·08-2·35] and 1·21 [1·15-1·27], respectively, for DCIS. In Asians, PRS_binary_ showed significant associations with BrCa (1·83 [1·64-2·05]) and DCIS (2·30 [1·88-2·83]). The GAIL_binary_ showed associations with BrCa (1·59 [1·31-1·92]) and DCIS (2·13 [1·51-3·00]) in Asians. The effect sizes for the associations between PRS_binary_, GAIL_binary_ and FH_binary_ were larger for younger Europeans than the older Europeans. In Asians, the same trend was observed for PRS_binary_ and BrCa, and FH_binary_ and DCIS.

**Table 2.** Association between high-risk criteria and case-control status (invasive breast cancer cases/ non-breast cancer controls), using univariate logistic regression. Analysis was repeated by age categories 30 to 49 years (n_European_=40,306, n_Asian_=8,456) and 50 to 80 years (n_European_=111,919, n_Asian_=9,113). PRS: 5-year absolute risk using polygenic risk score ≥1·66%. GAIL: 5-year absolute risk using the Gail model ≥1·66%. FH: having at least one first-degree family history of breast cancer. * High: individuals who were identified by any of the criteria were classified as “Yes”. OR: odds ratio, CI: confidence interval, P: p-value. 5-yr abs risk: 5-year absolute risk (continuous).

|  | All ages |  |  | Age 30 to 49 years |  |  | Age 50 to 80 years |  |  |
| --- | --- | --- | --- | --- | --- | --- | --- | --- | --- |
|  | OR (95% CI) | P | AUC (95% CI) | OR (95% CI) | P | AUC (95% CI) | OR (95% CI) | P | AUC (95% CI) |
| **European-ancestry, n=152,225** |  |  |  |  |  |  |  |  |  |
| 5-yr abs risk by PRS (continuous) | 1.97 (1.94 to 2.01) | <0.001 | 0.635 (0.632 to 0.638) | 2.51 (2.39 to 2.62) | <0.001 | 0.622 (0.617 to 0.628) | 2.06 (2.02 to 2.11) | <0.001 | 0.653 (0.650 to 0.656) |
| 5-yr abs risk by Gail (continuous) | 1.12 (1.11 to 1.14) | <0.001 | 0.492 (0.489 to 0.495) | 1.35 (1.29 to 1.40) | <0.001 | 0.493 (0.487 to 0.499) | 1.18 (1.16 to 1.19) | <0.001 | 0.517 (0.514 to 0.520) |
| PRS (Yes, ref=No) | 2.60 (2.52 to 2.69) | <0.001 | 0.552 (0.550 to 0.554) | 3.35 (3.02 to 3.70) | <0.001 | 0.530 (0.528 to 0.532) | 2.64 (2.55 to 2.74) | <0.001 | 0.562 (0.560 to 0.564) |
| Gail (Yes, ref=No) | 1.25 (1.22 to 1.28) | <0.001 | 0.522 (0.520 to 0.524) | 2.61 (2.36 to 2.88) | <0.001 | 0.523 (0.521 to 0.526) | 1.27 (1.24 to 1.30) | <0.001 | 0.527 (0.524 to 0.529) |
| FH (Yes, ref=No) | 1.56 (1.51 to 1.60) | <0.001 | 0.529 (0.527 to 0.530) | 1.87 (1.76 to 1.98) | <0.001 | 0.538 (0.535 to 0.542) | 1.47 (1.42 to 1.52) | <0.001 | 0.525 (0.523 to 0.527) |
| PRS/GAIL* (Yes, ref=No) | 1.62 (1.58 to 1.65) | <0.001 | 0.554 (0.552 to 0.557) | 2.98 (2.77 to 3.21) | <0.001 | 0.548 (0.545 to 0.551) | 1.67 (1.63 to 1.71) | <0.001 | 0.563 (0.560 to 0.565) |
| PRS/GAIL/FH* (Yes, ref=No) | 1.63 (1.60 to 1.67) | <0.001 | 0.558 (0.556 to 0.560) | 2.34 (2.22 to 2.47) | <0.001 | 0.566 (0.562 to 0.569) | 1.63 (1.59 to 1.67) | <0.001 | 0.560 (0.557 to 0.563) |
| **Asian-ancestry, n=17,569** |  |  |  |  |  |  |  |  |  |
| 5-yr abs risk by PRS (continuous) | 1.48 (1.41 to 1.56) | <0.001 | 0.564 (0.556 to 0.573) | 1.62 (1.47 to 1.78) | <0.001 | 0.551 (0.539 to 0.563) | 1.64 (1.53 to 1.75) | <0.001 | 0.600 (0.588 to 0.611) |
| 5-yr abs risk by Gail (continuous) | 1.19 (1.09 to 1.30) | <0.001 | 0.506 (0.497 to 0.514) | 0.94 (0.81 to 1.08) | 0.377 | 0.523 (0.511 to 0.535) | 1.82 (1.61 to 2.07) | <0.001 | 0.554 (0.543 to 0.566) |
| PRS (Yes, ref=No) | 1.83 (1.64 to 2.05) | <0.001 | 0.523 (0.519 to 0.527) | 2.24 (1.76 to 2.85) | <0.001 | 0.515 (0.511 to 0.519) | 1.89 (1.66 to 2.14) | <0.001 | 0.535 (0.528 to 0.542) |
| Gail (Yes, ref=No) | 1.59 (1.31 to 1.92) | <0.001 | 0.506 (0.503 to 0.508) | 1.34 (0.84 to 2.13) | 0.217 | 0.501 (0.499 to 0.503) | 1.77 (1.44 to 2.19) | <0.001 | 0.512 (0.507 to 0.516) |
| FH (Yes, ref=No) | 1.97 (1.76 to 2.19) | <0.001 | 0.527 (0.523 to 0.531) | 1.87 (1.59 to 2.19) | <0.001 | 0.524 (0.518 to 0.530) | 2.07 (1.78 to 2.41) | <0.001 | 0.530 (0.524 to 0.535) |
| PRS/GAIL* (Yes, ref=No) | 1.78 (1.61 to 1.96) | <0.001 | 0.527 (0.523 to 0.532) | 2.02 (1.63 to 2.50) | <0.001 | 0.516 (0.511 to 0.520) | 1.89 (1.69 to 2.12) | <0.001 | 0.543 (0.536 to 0.551) |
| PRS/GAIL/FH* (Yes, ref=No) | 1.91 (1.76 to 2.08) | <0.001 | 0.544 (0.539 to 0.550) | 1.95 (1.70 to 2.24) | <0.001 | 0.535 (0.528 to 0.542) | 2.01 (1.81 to 2.23) | <0.001 | 0.556 (0.548 to 0.564) |

**Table 3.** Association between high-risk criteria and case-control status (DCIS cases/ non-breast cancer controls), using univariate logistic regression. Analysis was repeated by age categories 30 to 49 years (n_European_=19,424, n_Asian_=4,322) and 50 to 80 years (n_European_= 58,740, n_Asian_=5,005). PRS: 5-year absolute risk using polygenic risk score ≥1·66%. GAIL: 5-year absolute risk using the Gail model ≥1·66%. FH: having at least one first-degree family history of breast cancer. * High: individuals who were identified by any of the three criteria (PRS, GAIL, FH) were classified as “Yes”. OR: odds ratio, CI: confidence interval, P: p-value. 5-yr abs risk: 5-year absolute risk (continuous).

|  | All ages |  |  | Age 30 to 49 years |  |  | Age 50 to 80 years |  |  |
| --- | --- | --- | --- | --- | --- | --- | --- | --- | --- |
|  | OR (95% CI) | P | AUC (95% CI) | OR (95% CI) | P | AUC (95% CI) | OR (95% CI) | P | AUC (95% CI) |
| **European-ancestry, n=78,164** |  |  |  |  |  |  |  |  |  |
| 5-yr abs risk by PRS (continuous) | 1.63 (1.59 to 1.68) | <0.001 | 0.626 (0.620 to 0.631) | 2.56 (2.37 to 2.78) | <0.001 | 0.657 (0.645 to 0.669) | 1.56 (1.51 to 1.61) | <0.001 | 0.620 (0.613 to 0.626) |
| 5-yr abs risk by Gail (continuous) | 1.23 (1.20 to 1.26) | <0.001 | 0.537 (0.531 to 0.543) | 2.28 (2.10 to 2.49) | <0.001 | 0.610 (0.597 to 0.622) | 1.19 (1.16 to 1.22) | <0.001 | 0.519 (0.512 to 0.526) |
| PRS (Yes, ref=No) | 2.21 (2.08 to 2.35) | <0.001 | 0.541 (0.537 to 0.544) | 3.30 (2.78 to 3.91) | <0.001 | 0.529 (0.523 to 0.535) | 2.11 (1.98 to 2.26) | <0.001 | 0.544 (0.539 to 0.549) |
| Gail (Yes, ref=No) | 1.21 (1.15 to 1.27) | <0.001 | 0.518 (0.513 to 0.523) | 3.35 (2.85 to 3.94) | <0.001 | 0.534 (0.527 to 0.540) | 1.12 (1.06 to 1.18) | <0.001 | 0.512 (0.506 to 0.518) |
| FH (Yes, ref=No) | 1.75 (1.65 to 1.85) | <0.001 | 0.537 (0.533 to 0.542) | 2.24 (2.00 to 2.50) | <0.001 | 0.553 (0.544 to 0.561) | 1.62 (1.52 to 1.72) | <0.001 | 0.532 (0.528 to 0.537) |
| PRS/GAIL* (Yes, ref=No) | 1.46 (1.40 to 1.53) | <0.001 | 0.542 (0.537 to 0.547) | 3.50 (3.09 to 3.96) | <0.001 | 0.559 (0.551 to 0.567) | 1.34 (1.28 to 1.41) | <0.001 | 0.536 (0.530 to 0.542) |
| PRS/GAIL/FH* (Yes, ref=No) | 1.52 (1.45 to 1.59) | <0.001 | 0.549 (0.544 to 0.554) | 2.74 (2.48 to 3.02) | <0.001 | 0.581 (0.572 to 0.591) | 1.36 (1.29 to 1.43) | <0.001 | 0.538 (0.531 to 0.544) |
| **Asian-ancestry, n=9,327** |  |  |  |  |  |  |  |  |  |
| 5-yr abs risk by PRS (continuous) | 1.67 (1.52 to 1.83) | <0.001 | 0.587 (0.566 to 0.607) | 1.70 (1.42 to 2.03) | <0.001 | 0.556 (0.528 to 0.584) | 1.89 (1.69 to 2.12) | <0.001 | 0.654 (0.628 to 0.680) |
| 5-yr abs risk by Gail (continuous) | 1.25 (1.03 to 1.52) | 0.023 | 0.507 (0.486 to 0.528) | 0.99 (0.71 to 1.38) | 0.93 | 0.533 (0.505 to 0.562) | 1.88 (1.46 to 2.41) | <0.001 | 0.542 (0.513 to 0.572) |
| PRS (Yes, ref=No) | 2.30 (1.88 to 2.83) | <0.001 | 0.535 (0.524 to 0.546) | 2.22 (1.43 to 3.43) | <0.001 | 0.514 (0.504 to 0.525) | 2.65 (2.09 to 3.36) | <0.001 | 0.561 (0.542 to 0.580) |
| Gail (Yes, ref=No) | 2.13 (1.51 to 3.00) | <0.001 | 0.511 (0.505 to 0.518) | 1.86 (0.81 to 4.26) | 0.144 | 0.503 (0.498 to 0.509) | 2.41 (1.64 to 3.53) | <0.001 | 0.521 (0.509 to 0.533) |
| FH (Yes, ref=No) | 2.76 (2.28 to 3.35) | <0.001 | 0.547 (0.535 to 0.558) | 2.89 (2.20 to 3.78) | <0.001 | 0.550 (0.533 to 0.566) | 2.64 (2.00 to 3.48) | <0.001 | 0.544 (0.527 to 0.560) |
| PRS/GAIL* (Yes, ref=No) | 2.23 (1.85 to 2.69) | <0.001 | 0.542 (0.530 to 0.554) | 2.09 (1.41 to 3.10) | <0.001 | 0.517 (0.505 to 0.528) | 2.63 (2.11 to 3.28) | <0.001 | 0.573 (0.553 to 0.593) |
| PRS/GAIL/FH* (Yes, ref=No) | 2.58 (2.21 to 3.02) | <0.001 | 0.571 (0.557 to 0.585) | 2.63 (2.06 to 3.37) | <0.001 | 0.557 (0.539 to 0.575) | 2.76 (2.24 to 3.39) | <0.001 | 0.589 (0.567 to 0.611) |

### Intersection of high-risk individuals identified by different risk factors

**Figure 1** illustrates the overlap of high-risk individuals identified by PRS_binary_, GAIL_binary_, and FH_binary_ across different ancestry and age groups. For young Europeans and all Asians, the proportion of high-risk individuals among BrCa (16-26%) and DCIS (20-30%) cases was about twice that of the controls (9-13%). In these groups, women were primarily classified as high-risk due to FH_binary_ and PRS_binary_. Less than 7%, 10%, and 3% of the BrCa, DCIS, and controls, respectively, were classified as high-risk by more than one criterion (i.e. PRS_binary_, GAIL_binary_, or FH_binary_). PRS_binary_ uniquely identified 4-7% of young BrCa and DCIS cases as high-risk, and 10-18% of older BrCa and DCIS cases as high-risk. FH_binary_ uniquely identified 9-11% of young BrCa and DCIS cases as high-risk, and 2-5% of older BrCa and DCIS cases as high-risk. The proportion of young Europeans (aged 30-49) uniquely identified by GAIL_binary_ to be at high-risk is 1%. Among Asians, all individuals classified as GAIL_binary_ high-risk were also positive for FH (i.e. 0% uniquely called by GAIL_binary_). Among the older Europeans, 40% of the controls were identified as high-risk, compared to 52% of BrCa cases and 47% of DCIS cases.

**Figure 1.**
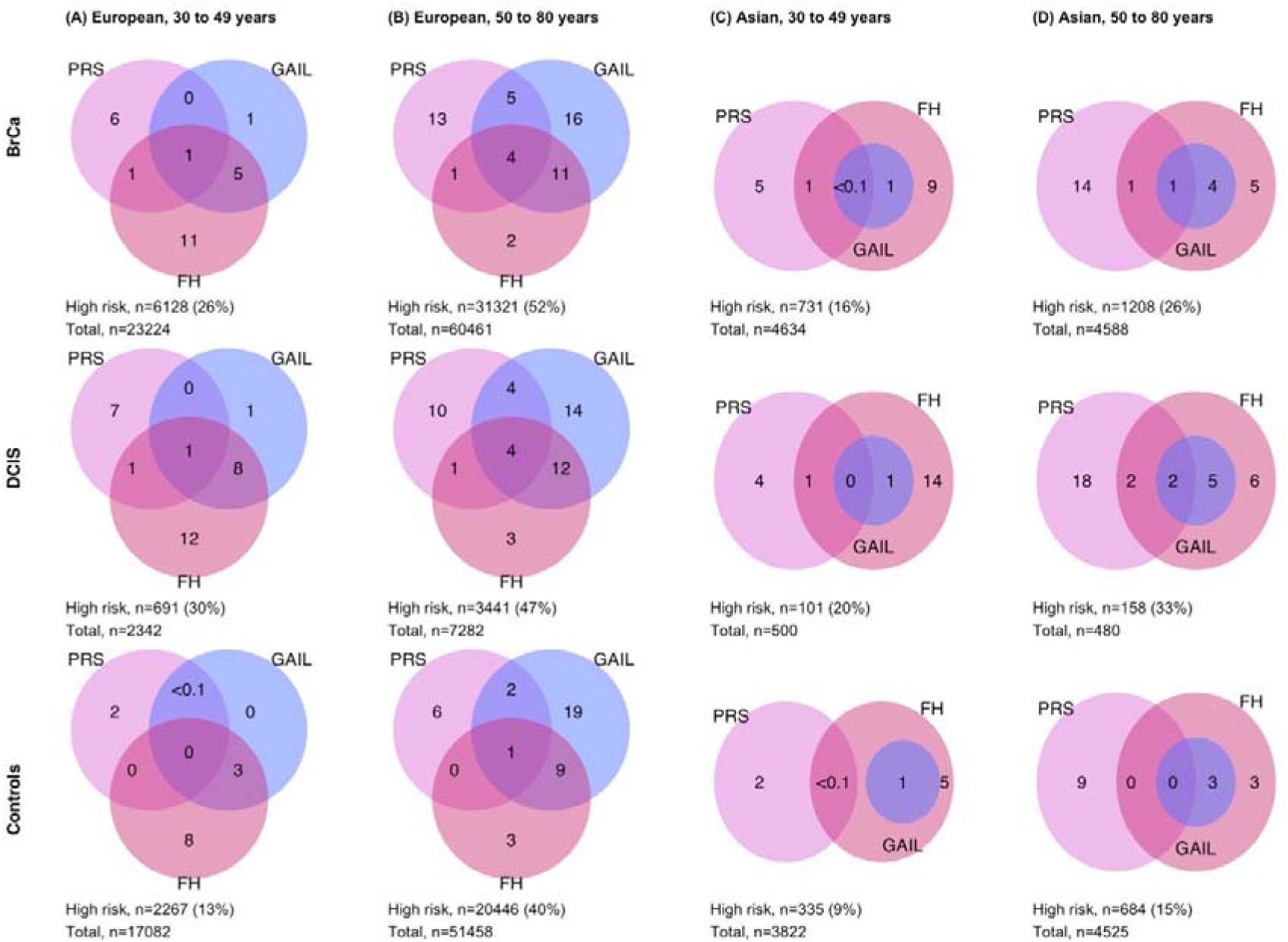
Venn diagram depicting the overlaps between individuals identified as high risk by the three criteria. Breast cancer polygenic risk score (PRS, 5-year absolute risk using polygenic risk score ≥1·66%), the Gail model (GAIL, 5-year absolute risk using the Gail model ≥1·66%), and family history (FH, having at least one first-degree family history of breast cancer). European: women of European ancestry; Asian: women of Asian ancestry.

### Breast cancer predisposition genes (PTV_binary_) and common variants (PRS_binary_) identified different high-risk individuals

In the subgroup of individuals with target-enriched sequencing data, the proportion of women identified to be at high risk by both PRS_binary_ and PTV_binary_ was limited (0·6% of European and 0·1% of Asian) compared to PTV_binary_ alone (Europeans: 4·6%; Asians: 4·4%) and PRS_binary_ alone (Europeans: 13·9%; Asians: 8·5%) (**Table 4).** There were more older women than younger women at high risk due to PRS (2·3x in Europeans; 3·8x in Asians). Conversely, there were more younger women than older women at high risk due to PTV (1.9x in Europeans; 1.5x in Asians).

**Table 4.**
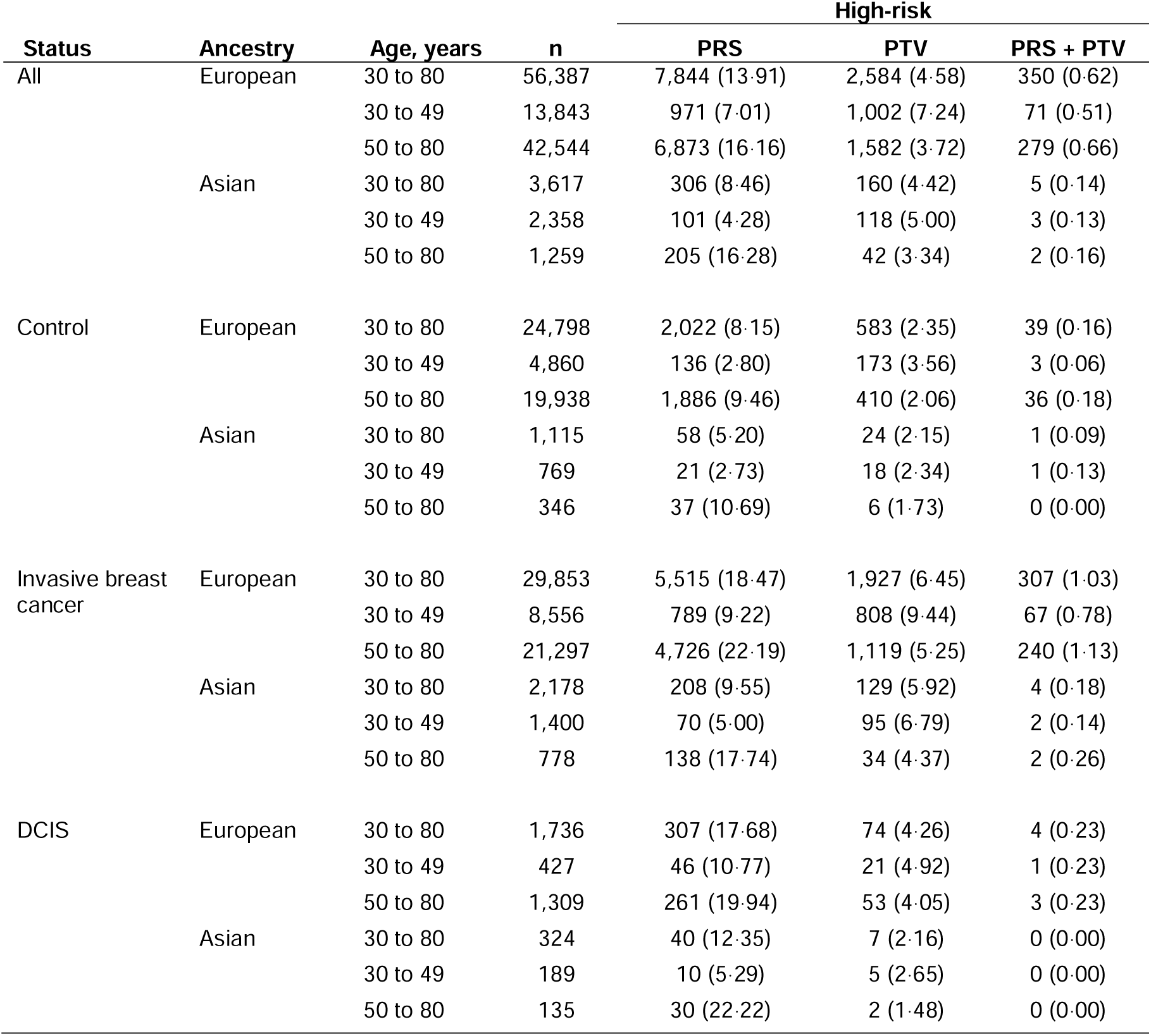
Overlap of individuals with 5-year absolute risk by polygenic risk score ≥1·66% (PRS) and carriers of protein-truncating variants in at least one of nine breast cancer predisposition genes (i.e. PTVs in *ATM, BRCA1, BRCA2, CHEK2, PALB2, BARD1, RAD51C, RAD51D,* or *TP53*).

### Country-specific differences in high-risk individuals identified by PRS and GAIL

**Figure 2** shows the breakdown of BrCa/DCIS cases and controls identified to be at high risk by PRS_binary_ and GAIL_binary_ by country and age. Generally, both PRS_binary_ and GAIL_binary_ identified a higher proportion (%) of high-risk BrCa/DCIS individuals in the European-ancestry populations (median [IQR], PRS_binary-young_: 5 [3-9], PRS_binary-old_: 20 [10-23], GAIL_binary-young_: 3 [2-7], GAIL_binary-old_: 30 [24-39]) than the Asian countries (PRS_binary-young_: 4 [2-5], PRS_binary-old_: 15 [10-16], GAIL_binary-young_: <1 [<1 to 2], GAIL_binary-old_: 4 [2-10]).

**Figure 2.**
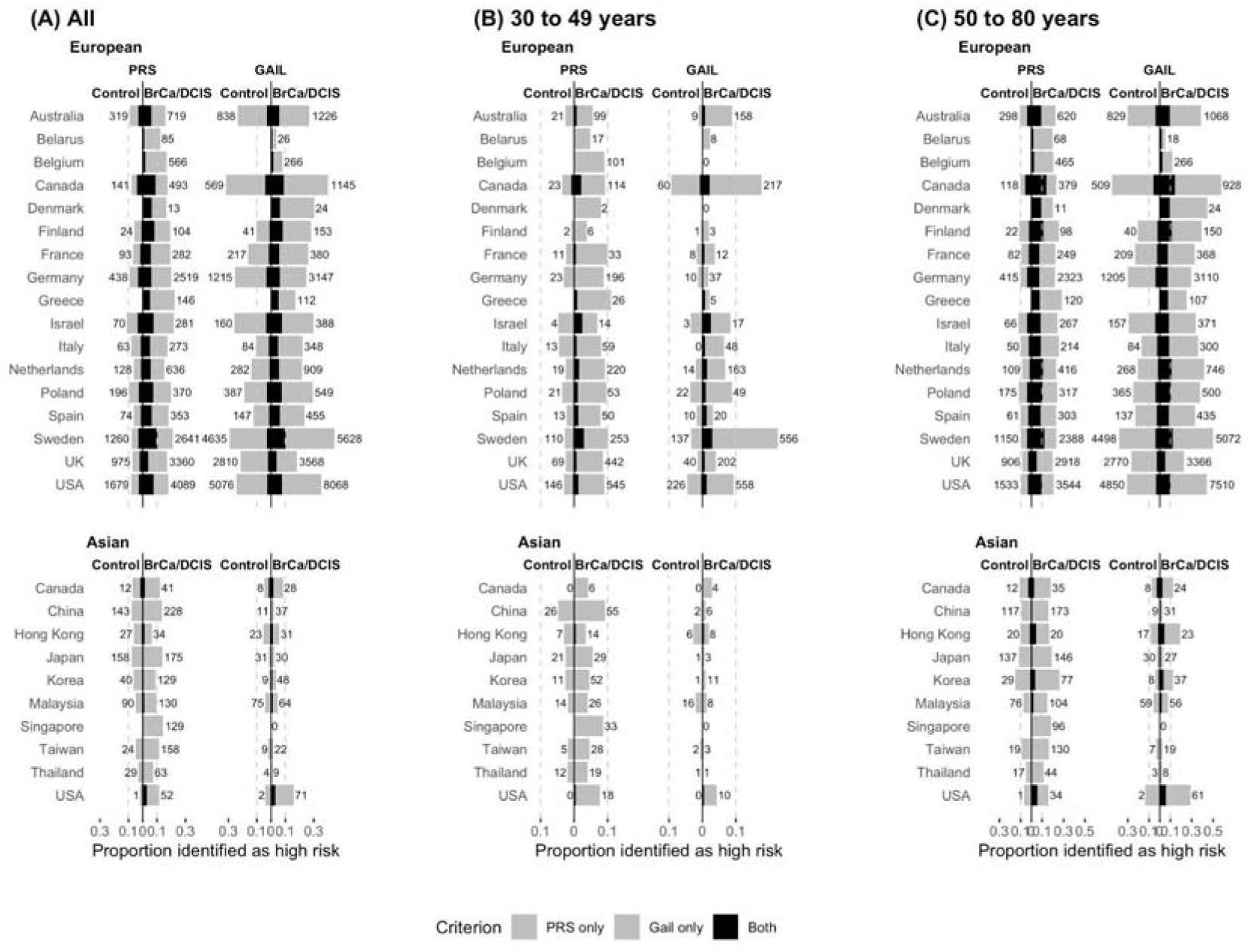
The proportion of individuals identified as at high risk by the breast cancer polygenic risk score (PRS) and the Gail model (GAIL), by country and age. The proportion of individuals identified as high risk by both criteria is indicated in grey. PRS: 5-year absolute risk using polygenic risk score ≥1·66%. GAIL: 5-year absolute risk using the Gail model ≥1·66%. Numbers adjacent to the bars represent the number of high-risk individuals identified by respective risk tools. European: women of European ancestry; Asian: women of Asian ancestry.

### Factors influencing the performance of the Gail model across different demographics

**Figure 2** shows that both PRS_binary_ and GAIL_binary_ identified a higher proportion of high-risk individuals in European-ancestry compared to Asian-ancestry populations, with variations in performance by age. By studying the factors that influence the Gail model’s performance across different demographics, we can better understand what drives the model’s effectiveness in various populations. In **Figure 3A** and **Figure 3B**, we show that in Europeans, incorporating both family history (number of first-degree relatives with breast cancer) and prior breast biopsies was sufficient to achieve the highest AUC. For younger Asians, the key factors affecting model performance are age at menarche and the number of prior biopsies, with higher discriminatory ability observed in models that did not include family history (**Figure 3C**). In older Asians, the model’s performance was not significantly different between those that included both age at first live birth and family history (**Figure 3D**). This indicates that, while the Gail model’s performance for Europeans is primarily influenced by the number of first-degree relatives with breast cancer and prior biopsies, younger Asians are more influenced by age at first menarche, and for older Asians, both age at first live birth and family history are important.

**Figure 3.**
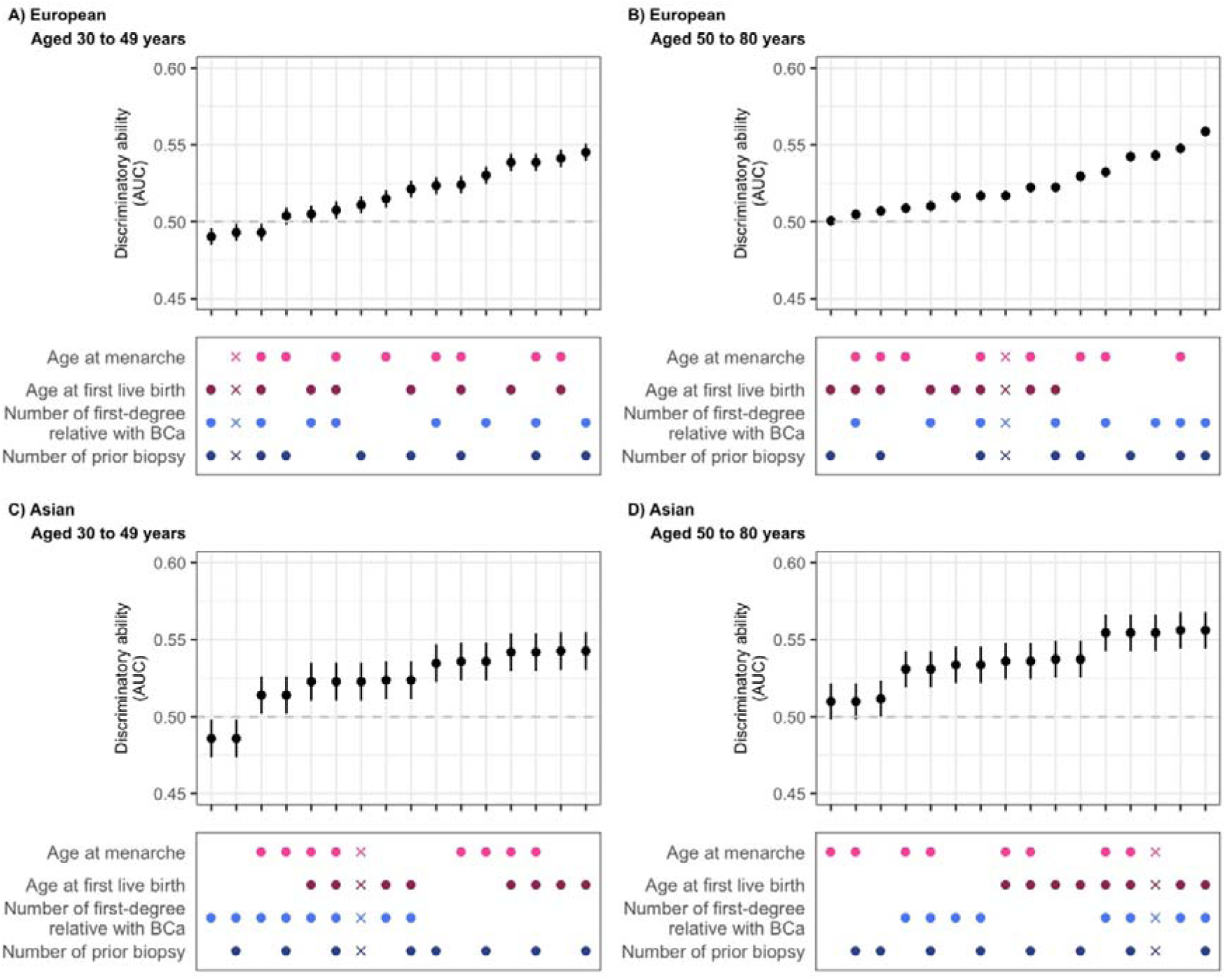
Discriminatory ability of risk factor combinations in the Gail model. The five-year absolute risk was calculated using the R package “BCRA” and used to predict the invasive breast cancer case-control status of the individuals. Dots represent risk factors included in the model, and crosses indicate the model with all risk factors with the addition of atypical hyperplasia. European: women of European ancestry; Asian: women of Asian ancestry.

Applying the high-risk criterion of ≥1·66% 5-year absolute risk resulted in changes in the order of the models regarding their discriminatory ability (AUC) (**Supplementary Figure 4**). However, the AUCs of the models were small (highest AUC [95%CI]: 0·529 [0·526-0·531]) and not appreciably different (within 0·03 difference in AUCs. Excluding models with confidence intervals including 0·5, lowest AUC: 0·501 [0·500-0·501]) (**Supplementary Table 2**).

## Discussion

We evaluated the performance of different risk stratifiers, including PRS, GAIL, and FH, in identifying high-risk individuals for BrCa and DCIS across various demographics and to understand the overlap and unique contributions of these models in different populations. The association between different risk stratifiers (PRS, GAIL, and FH) with BrCa and DCIS varies by ancestry and age. PRS demonstrated superior discrimination compared to the Gail model for predicting both BrCa and DCIS in European- and Asian-ancestry populations. Specifically, the 5-year absolute risk from PRS showed higher AUC values than the Gail model for both conditions. In Europeans, PRS and GAIL showed significant odds ratios for identifying high-risk individuals, with larger effect sizes observed in younger populations. In Asians, PRS and GAIL also showed significant associations. The overlap of high-risk individuals identified by PRS, GAIL, and FH revealed that PRS and FH were primary contributors to high-risk classification, particularly in young Europeans and all Asians. PRS uniquely identified a notable percentage of high-risk individuals that were missed by GAIL and FH, while GAIL identified a significant portion in older Europeans. Additionally, target-enriched sequencing data showed that high-risk individuals identified by both PRS and predisposition genes (PTV) were limited, with PRS alone identifying a larger proportion of high-risk individuals compared to PTV alone. Country-specific analysis indicated that both PRS and GAIL identified a higher proportion of high-risk individuals in European-ancestry populations compared to Asian-ancestry populations, with greater variability observed for GAIL.

Given the complexity and multifactorial nature of breast cancer, relying on a single risk factor or model may not sufficiently capture all high-risk individuals.^6,12^ The analysis of the intersection between high-risk individuals identified by PRS, GAIL, and FH reveals important insights into how these risk stratifiers overlap and uniquely contribute to risk assessment. Our results derived from the analysis of 180,398 woman across diverse ancestries corroborate previous findings that report a limited overlap in the high-risk individuals identified by different risk predictors.^6,12^ The unique contribution of PRS is particularly notable. Traditional risk models like the Gail model are less accurate in younger populations. These models often rely on risk factors which may not fully capture the risk in younger women. Younger women may not have a significant personal or reproductive history, making genetic information from PRS and specific gene mutations particularly valuable for risk assessment.^6,12^ No single model is suitable for every subgroup within the general population. The limited overlap and the unique contributions of each risk stratifier suggest that using a combination of these tools could provide a more comprehensive risk assessment, capturing high-risk individuals that might be missed by any single model. Comprehensive risk models such as BOADICEA improve prediction, however they can be challenging to implement at the general population level.^18^ In addition, calibration and validation for populations not used in the model’s development need to be done.

The evaluation of country-specific differences in high-risk identification by PRS and GAIL shows that European-ancestry populations generally had higher proportions of predicted high-risk individuals compared to Asian-ancestry populations. This is expected, as two risk predictors utilize breast cancer incidence rates that were higher in Europeans (i.e. the “White” used to develop the Gail model) than in Asians (i.e. the “Chinese”).^15^ The variability in the performance of the GAIL model across different countries, with larger standard deviations compared to PRS, suggests that GAIL’s effectiveness may be more influenced by regional factors, such as differences in reproductive factors, lifestyle and healthcare practices.^11,25^ The analysis of factors influencing the performance of the Gail model reveals differences in its effectiveness based on age and demographic factors. For Europeans, incorporating family history and prior breast biopsies achieved the highest AUC, emphasizing the importance of these factors in risk prediction. In younger Asians, age at menarche and the number of prior biopsies were more influential, with models excluding family history showing better performance. While there are likely regional differences in genetics, lifestyle, and healthcare practices, the Gail model may be compounded by variations in data quality and recall.^25,26^

The current guidelines for breast cancer screening are based on sex and age. The recommendations typically advocate biennial mammography for women aged 50 to 69 or 70 years.^3,27^ Previously, the US Preventive Services Task Force (USPSTF) advised that the decision to begin biennial screening before age 50 should be personalized, considering the patient’s values regarding specific benefits and harms. It is unclear if clinicians are provided with directives on the specific topics to discuss with patients regarding screening suitability. For age groups where the evidence for mammography is less definitive, integrating comprehensive risk stratification into discussions about screening would ensure that recommendations are as relevant as possible. However, in Apr 2024, the USPSTF published in its Final Recommendation Statement biennial mammogram screenings for all women aged 40 to 74 years to detect early-stage cancer (accessed Jul 23, 2024).^28^ While this earlier screening will benefit many, it also raises concerns about over-screening and its potential consequences. Risk stratification could enhance the effectiveness of the new USPSTF guidelines by targeting screening efforts more precisely. By incorporating complementary individual risk factors, healthcare providers can better identify those who are genuinely at higher risk for breast cancer. As a result, risk stratification can help balance the benefits of early detection with the potential drawbacks of excessive screening.

Our study uses one of the largest breast cancer association study datasets, providing high statistical power for comprehensive risk factor analyses and diverse population coverage that includes both European and Asian ancestries. The study’s multi-center scope allowed for the comparison of risk models across different countries. However, differences in study design, data collection methods, and risk factor definitions across included studies may have introduced variability and affected the consistency of results. Variations in the time of data collection and changes in clinical practices over time could affect the comparability of data across studies. Combining different studies introduced gaps in data (i.e. missingness) for some risk factors, and exclusions of certain studies affected the generalizability of our findings for those regions. In addition, while the study covers women of European and Asian ancestries, it does not represent all global populations. Regional variations in absolute risk—due to genetic differences, varying gene predispositions, lifestyle factors, and healthcare access—also impact the applicability of risk models to other settings or demographics.^11,29^ Not all known breast cancer risk factors were considered in our analyses. Examples of other risk factors include mammographic density, physical activity, alcohol use, and smoking.^18,30^

Overall, while PRS shows consistent superiority in breast cancer risk stratification across demographics, the complementary use of the Gail model and family history can enhance the overall risk assessment process. Integrating and calibrating these models for different ethnic populations, along with understanding their unique contributions and limitations, can lead to more precise identification of high-risk individuals who would otherwise be missed.^13^ Ideally, evaluating individual breast cancer risk could lead to more precise and cost-effective early detection by tailoring screening approaches to risk levels. However, despite advancements in risk assessment, it remains important to adhere to the established consensus guidelines for minimum mammography screening, as set by nationally recognized organizations with expertise in screening methodology.

## DECLARATIONS

### Ethics approval

This study was approved by the A*STAR Institutional Review Board (reference number: 2022-041).

### Consent to participate

Informed consent was obtained by the individual studies that contributed to BCAC.

### Availability of data and materials

The data used in our analyses are available upon reasonable request through BCAC, subject to data access committee approval.

## Acknowledgements

This study is funded by the Agency for Science, Technology and Research (A*STAR) and PRECISION Health Research, Singapore (PRECISE). The breast cancer genome-wide association analyses in BCAC were supported by the Government of Canada through Genome Canada and the Canadian Institutes of Health Research, the ‘Ministère de l’Économie, de la Science et de l’Innovation du Québec’ through Genome Québec and grant PSR-SIIRI-701, The National Institutes of Health (U19 CA148065, X01HG007492), Cancer Research UK (C1287/A10118, C1287/A16563, C1287/A10710), and The European Union (HEALTH-F2-2009-223175 and H2020 633784 and 634935). All studies and funders are listed in **Additional Materials (BCAC Funding and Acknowledgments)**.

## Author contributions

Conception and design of the study: JL; Acquisition of data: All; Analysis and interpretation of data: PJH, MHG, CKYL, JL; Drafting of the manuscript or revising it for important content: JL, PJH, CKYL; Final approval of the version submitted for publication: All.

