## Supplementary Materials for "Overlap of high-risk individuals across family history, genetic & non-genetic breast cancer risk models: Analysis of 180,398 women from European & Asian ancestries"

### **Supplementary Tables in Main Text**

#### Supplementary Table 1. List of studies included.

| **Study** | **Acronym** | **Country** | **Type of control** | **Identifier for within study strata** | **Identifier for within study strata** | **Specify (other)** | **n** | **DOI** |
| --- | --- | --- | --- | --- | --- | --- | --- | --- |
| The Two Sister Study | 2SISTER | USA | Not applicable (for cases) | Not applicable | Other | Population based familial | 1067 | 10.1093/jnci/djs255 |
| Australian Breast Cancer Family Study | ABCFS | Australia | Population-based | Not applicable | Control |  | 2065 | 10.1093/jnci/95.6.448 |
| Amsterdam Breast Cancer Study | ABCS | Netherlands | Not applicable (for cases) | ABCS-2 | Sporadic (population or hospital based) |  | 2477 | 10.1200/JCO.2006.06.3024 |
| Australian Breast Cancer Tissue Bank | ABCTB | Australia | Not applicable (for cases) | Not applicable | Sporadic (population or hospital based) |  | 856 | 10.5334/ojb.aa |
| Asia Cancer Program | ACP | Thailand | Hospital-based | Not applicable | Control |  | 1806 | 10.2188/jea.JE20170045 |
|  | AHS | USA | Nested case-control | Don't Know | Control |  | 1624 | 10.1007/s10552-019-01140-y |
| Bavarian Breast Cancer Cases and Controls, | BBCC | Germany | Hospital-based | BBCC2 | Control |  | 1495 | 10.1007/s00432-008-0355-9 |
| British Breast Cancer Study, | BBCS | UK | Not applicable (for cases) | NCRN | Other |  | 106 | 10.1093/jnci/djj268 |
| Breast Cancer Employment and Environment Study | BCEES | Australia | Population-based | Not applicable | Control |  | 1615 | 10.1038/bjc.2013.544 |
| Philadelphia Breast Cancer Family Registry, | BCFR-PA | USA | Don't Know | Don't Know | Familial (clinical genetic centre based) |  | 66 | 10.1186/bcr801 |
| Breast Cancer in Northern Israel Study, | BCINIS | Israel | Population-based | Not applicable | Control |  | 1920 | 10.1200/JCO.2010.28.1113 |
| Breast Oncology Galicia Network, | BREOGAN | Spain | Population-based | Don't Know | Control |  | 2494 | 10.3892/or.17.5.1109 |
| Canadian Breast Cancer Study- | CBCS | Canada | Population-based | Don't Know | Control |  | 1995 | 10.1016/j.canep.2013.04.006 |
| Crete Cancer Genetics Program | CCGP | Greece | Not applicable (for cases) | Not applicable | Sporadic (population or hospital based) |  | 658 | 10.1016/j.annonc.2022.03.166 |
| CECILE Breast Cancer Study | CECILE | France | Population-based | Not applicable | Control |  | 2010 | 10.1002/ajim.20952 |
| Copenhagen General Population Study | CGPS | Denmark | Not applicable (for cases) | Copenhagen hospital | Sporadic (population or hospital based) |  | 79 | 10.1200/JCO.2005.05. |
| Cancer Prevention Study-II Nutrition Cohort | CPSII | USA | Not applicable (for cases) | NUTRITION COHORT | Sporadic (population or hospital based) |  | 6240 | 10.1002/cncr.10197 |
| California Teachers Study | CTS | USA | Don't Know | Not applicable | Control |  | 1803 | 10.1023/a:1019552126105 |
| DietCompLyf Breast Cancer Survival Study | DIETCOMPLYF | UK | Not applicable (for cases) | Not applicable | Sporadic (population or hospital based) |  | 711 | 10.1016/j.maturitas.2013.03.018 |
| European Prospective Investigation Into Cancer and Nutrition | EPIC | France | Nested case-control | EPIC France | Control |  | 800 | 10.1079/PHN2002394 |
| European Prospective Investigation Into Cancer and Nutrition | EPIC | Germany | Nested case-control | EPIC Germany | Control |  | 1307 | 10.1079/PHN2002394 |
| European Prospective Investigation Into Cancer and Nutrition | EPIC | Italy | Nested case-control | EPIC Italy | Control |  | 1601 | 10.1079/PHN2002394 |
| European Prospective Investigation Into Cancer and Nutrition | EPIC | Netherlands | Not applicable (for cases) | EPIC The Netherlands | Sporadic (population or hospital based) |  | 1364 | 10.1079/PHN2002394 |
| European Prospective Investigation Into Cancer and Nutrition | EPIC | Spain | Nested case-control | EPIC Spain | Control |  | 643 | 10.1079/PHN2002394 |
| European Prospective Investigation Into Cancer and Nutrition | EPIC | UK | Nested case-control | EPIC United Kingdom | Control |  | 1346 | 10.1079/PHN2002394 |
| ESTHER Breast Cancer Study | ESTHER | Germany | Population-based | e1 | Control |  | 984 | 10.1371/journal.pone.0002656 |
|  | FHRISK | UK | Not applicable (for cases) | Not applicable | Familial (clinical genetic centre based) |  | 748 | 10.1007/s10549-021-06333-1 |
| Gene Environment Interaction and Breast Cancer in Germany, | GENICA | Germany | Not applicable (for cases) | Not applicable | Sporadic (population or hospital based) |  | 1626 | 10.1007/s10654-005-0032-0 |
| Genetic Epidemiology Study of Breast Cancer by Age | GESBC | Germany | Not applicable (for cases) | Not applicable | Sporadic (population or hospital based) |  | 504 | 10.1023/a:1008907901087 |
| Hannover Breast Cancer Study | HABCS | Germany | Not applicable (for cases) | HaBCS1 | Sporadic (population or hospital based) |  | 896 | 10.1038/nature05887 |
| Hospital-based Epidemiologic Research Program at Aichi Cancer Center | HERPACC | Japan | Hospital-based | Not applicable | Control |  | 2543 | <https://pubmed.ncbi.nlm.nih.gov/12718687/> |
| Hong Kong Breast Cancer Study, | HKBCS | Hong Kong | Population-based | Not applicable | Control |  | 953 | <https://rfs2.healthbureau.gov.hk/app/fundedsearch/projectdetail.xhtml?id=1903> |
| Hannover-Minsk Breast Cancer Study | HMBCS | Belarus | Not applicable (for cases) | Not applicable | Sporadic (population or hospital based) |  | 689 | 10.1007/s10549-008-0189-9 |
|  | ICICLE | UK | Not applicable (for cases) | Not applicable | Sporadic (population or hospital based) |  | 2823 | 10.1056/NEJMoa1913948 |
| Karolinska Breast Cancer Study, | KARBAC | Sweden | Not applicable (for cases) | Not applicable | Sporadic (population or hospital based) |  | 760 | 10.1089/gte.2004.8.127 |
| Karolinska Mammography Project for Risk Prediction of Breast Cancer | KARMA | Sweden | Population-based | KARMA | Control |  | 10100 | 10.1093/ije/dyw357 |
| Kuopio Breast Cancer Project, | KBCP | Finland | Not applicable (for cases) | Not applicable | Sporadic (population or hospital based) |  | 945 | PMID: 15668479 |
| Kathleen Cuningham Foundation Consortium for Research into Familial Aspects of Breast Cancer (kConFab)/Australian Ovarian Cancer Study | KCONFAB/AOCS | Australia | Not applicable (for cases) | kConFab | Familial (clinical genetic centre based) |  | 1401 | 10.1158/1055-9965.EPI-07-0542 |
| Korean Hereditary Breast Cancer Study | KOHBRA | Korea | National 43 centers (for cases) | KOHBRA | Familial (National 43 genetic centre based) or Controls (population or center based control) | Controls includes carriers' family members or normal control from general population | 1936 | 10.1016/j.clon.2010.11.007 |
| Leuven Multidisciplinary Breast Centre, | LMBC | Belgium | Not applicable (for cases) | retrospective | Sporadic (population or hospital based) |  | 3364 | 10.1007/s10549-008-0189-9 |
| Mammary Carcinoma Risk Factor Investigation | MARIE | Germany | Population-based | Not applicable | Control |  | 3867 | 10.1002/ijc.23655 |
| Milan Breast Cancer Study Group, | MBCSG | Italy | Not applicable (for cases) | IEO | Familial (clinical genetic centre based) |  | 757 | 10.1093/carcin/bgm290 |
| Mayo Clinic Breast Cancer Study | MCBCS | USA | Hospital-based | Not applicable | Control |  | 4268 | 10.1158/1055-9965.EPI-06-0781 |
| Melbourne Collaborative Cohort Study | MCCS | Australia | Nested case-control | Not applicable | Control |  | 2163 | PMID: 12484128 |
| Multi-ethnic Cohort | MEC | USA | Not applicable (for cases) | Don't Know | Sporadic (population or hospital based) |  | 1576 | 10.1038/nrc1389 |
| Melanoma Inquiry of Southern Sweden | MISS | Sweden | Not applicable (for cases) | Don't Know | Sporadic (population or hospital based) |  | 2136 |  |
| Mayo Mammography Health Study | MMHS | USA | Nested case-control | Not applicable | Control |  | 1954 | 10.1158/1538-7445.AM2011-3716 |
|  | MSKCC | USA | Not applicable (for cases) | TNBCC | Familial (clinical genetic centre based) |  | 135 | 10.1158/0008-5472.CAN-11-3364 |
| Malaysian Breast Cancer Genetic Study | MYBRCA | Malaysia | Not applicable (for cases) | Not applicable | Sporadic (population or hospital based) |  | 3152 | 10.1371/journal.pone.0203469 |
| Nashville Breast Health Study | NBHS | USA | Not applicable (for cases) | Not applicable | Sporadic (population or hospital based) |  | 1387 | [10.1007/s10549-011-1538-7](https://doi.org/10.1007%2Fs10549-011-1538-7) |
| Northern California Breast Cancer Family Registry | NC-BCFR | USA | Not applicable (for cases) | Not applicable | Other | Population based familial | 1392 | 10.1007/s10552-019-01154-6. |
|  | NCBCS | USA | Not applicable (for cases) | Not applicable | Sporadic (population or hospital based) |  | 3369 | 10.1007/BF00694745 |
| Nagano Breast Cancer Study | NGOBCS | Japan | Hospital-based | Not applicable | Control |  | 731 | 10.1038/bjc.2014.223 |
| Nurses Health Study | NHS | USA | Nested case-control | Not applicable | Control |  | 3075 | 10.1089/jwh.1997.6.49 |
| Nurses Health Study | NHS2 | USA | Nested case-control | Not applicable | Control |  | 3423 |  |
| Ontario Familial Breast Cancer Registry | OFBCR | Canada | Not applicable (for cases) | Not applicable | Control |  | 3156 | 10.1186/bcr801 |
| Leiden University Medical Centre Breast Cancer Study, | ORIGO | Netherlands | Not applicable (for cases) | Not applicable | Sporadic (population or hospital based) |  | 1351 | 10.1136/jmg.2004.019737 |
| NCI Polish Breast Cancer Study | PBCS | Poland | Not applicable (for cases) | Not applicable | Sporadic (population or hospital based) |  | 3953 | 10.1038/sj.bjc.6603207 |
|  | PKARMA | Sweden | Not applicable (for cases) | PKARMA | Sporadic (population or hospital based) |  | 10923 | 10.1038/ng.2563 |
| The Prostate, Lung, Colorectal and Ovarian (PLCO) Cancer Screening Trial | PLCO | USA | Not applicable (for cases) | Don't Know | Sporadic (population or hospital based) |  | 4808 | 10.1186/1471-2407-9-84 |
|  | PREFACE | Germany | Not applicable (for cases) | Not applicable | Sporadic (population or hospital based) |  | 2842 | 10.1055/a-2238-3153 |
|  | PROCAS | UK | Population-based | Not applicable | Control |  | 2123 | https://www.ncbi.nlm.nih.gov/books/NBK379493/ |
| Rotterdam Breast Cancer Study | RBCS | Netherlands | Not applicable (for cases) | Not applicable | Familial (clinical genetic centre based) |  | 1038 | 10.1007/s10549-010-1080-z |
|  | SASBAC | Sweden | Population-based | Not applicable | Control |  | 2502 | 10.1186/bcr811 |
| Shanghai Breast Cancer Genetic Study | SBCGS | China | Population-based | Don't Know | Control |  | 3511 | 10.1038/ng.318 |
|  | SBCS | UK | Not applicable (for cases) | Not applicable | Sporadic (population or hospital based) |  | 1743 | 10.1093/jnci/dji001 |
| Study of Epidemiology and Risk factors in Cancer Heredity | SEARCH | UK | Not applicable (for cases) | SEARCH | Sporadic (population or hospital based) |  | 21332 | 10.1038/nature05887 |
| Seoul Breast Cancer Study, | SEBCS | Korea | Not applicable (for cases) | Not applicable | Sporadic (population or hospital based) |  | 135 | 10.1186/1471-2407-5-143 |
| Singapore Breast Cancer Cohort | SGBCC | Singapore | Not applicable (for cases) | SBCCP | Sporadic (population or hospital based) |  | 894 | 10.1371/journal.pone.0250102 |
| The Sister Study | SISTER | USA | Not applicable (for cases) | Not applicable | Familial (clinical genetic centre based) |  | 3548 | 10.1289/EHP1923 |
| Städtisches Klinikum Karlsruhe Deutsches Krebsforschungszentrum Study | SKKDKFZS | Germany | Not applicable (for cases) | Not applicable | Sporadic (population or hospital based) |  | 1174 | 10.1016/j.ejca.2005.04.049 |
| Swedish Mammography Cohort | SMC | Sweden | Nested case-control | Not applicable | Control |  | 2086 | https://snd.se/en/catalogue/dataset/ext0018-5 |
|  | SUCCESSC | Germany | Not applicable (for cases) | NA | Familial (clinical genetic centre based) |  | 2816 | 10.1159/000322677 |
| Taiwanese Breast Cancer Study, | TWBCS | Taiwan | Not applicable (for cases) | Not applicable | Sporadic (population or hospital based) |  | 1889 | 10.1093/hmg/ddn429 |
|  | UBCS | USA | Not applicable (for cases) | Other | Sporadic (population or hospital based) |  | 645 | 10.3390/nu11081883 |
| UCI Breast Cancer Study, | UCIBCS | USA | Not applicable (for cases) | Not applicable | Sporadic (population or hospital based) |  | 726 | 10.1093/hmg/ddn429 |
| UK Breakthrough Generations Study | UKBGS | UK | Nested case-control | Not applicable | Control |  | 2057 | 10.1038/bjc.2011.337 |
|  | USRT | USA | Not applicable (for cases) | Don't Know | Sporadic (population or hospital based) |  | 3341 | 10.1002/1097-0142(19920115)69:2<586::aid-cncr2820690251>3.0.co;2-3. |

#### Supplementary Table 2. Discriminatory ability of combinations of risk factors included in the Gail model. Risk factors not listed were set to unknown for the 5-year absolute risk to be calculated for the combination specified. Agemen: age of menarche, Age1st: age at first live birth, N_Rel: number of family (first-degree) history of breast cancer, N_Biop: number of breast biopsies. *Full_model includes atypical hyperplasia

|  | **AUC (95% confidence interval)** | |
| --- | --- | --- |
|  | **5-year absolute risk from R package "BRCA" (abs_risk)** | |
| **Risk factor combinations** | **Continuous** | **High risk (binary, abs_risk>=1.66%)** |
| **European, aged 30 to 49 years** |  |  |
| Age1st | 0.539 (0.533 to 0.544) |  |
| Age1st + N_Biop | 0.521 (0.516 to 0.527) | 0.507 (0.507 to 0.508) |
| Age1st + N_Rels | 0.505 (0.499 to 0.511) | 0.508 (0.507 to 0.510) |
| Age1st + N_Rels + N_Biop | 0.490 (0.485 to 0.496) | 0.518 (0.516 to 0.520) |
| AgeMen | 0.515 (0.509 to 0.521) |  |
| AgeMen + Age1st | 0.541 (0.536 to 0.547) |  |
| AgeMen + Age1st + N_Biop | 0.524 (0.518 to 0.530) | 0.509 (0.508 to 0.510) |
| AgeMen + Age1st + N_Rels | 0.508 (0.502 to 0.513) | 0.514 (0.512 to 0.516) |
| AgeMen + Age1st + N_Rels + N_Biop | 0.493 (0.487 to 0.499) | 0.523 (0.521 to 0.526) |
| AgeMen + N_Biop | 0.504 (0.498 to 0.510) | 0.501 (0.500 to 0.501) |
| AgeMen + N_Rels | 0.524 (0.518 to 0.529) | 0.511 (0.510 to 0.513) |
| AgeMen + N_Rels + N_Biop | 0.539 (0.533 to 0.544) | 0.515 (0.514 to 0.517) |
| Full_model* | 0.493 (0.487 to 0.499) | 0.523 (0.521 to 0.526) |
| N_Biop | 0.511 (0.505 to 0.517) |  |
| N_Rels | 0.530 (0.525 to 0.536) | 0.507 (0.505 to 0.508) |
| N_Rels + N_Biop | 0.545 (0.540 to 0.551) | 0.511 (0.510 to 0.512) |
| **European, aged 50 to 80 years** |  |  |
| Age1st | 0.516 (0.513 to 0.520) | 0.501 (0.499 to 0.504) |
| Age1st + N_Biop | 0.501 (0.497 to 0.504) | 0.511 (0.509 to 0.514) |
| Age1st + N_Rels | 0.510 (0.507 to 0.514) | 0.521 (0.518 to 0.523) |
| Age1st + N_Rels + N_Biop | 0.522 (0.519 to 0.526) | 0.529 (0.526 to 0.531) |
| AgeMen | 0.509 (0.505 to 0.512) |  |
| AgeMen + Age1st | 0.522 (0.519 to 0.526) | 0.500 (0.498 to 0.503) |
| AgeMen + Age1st + N_Biop | 0.507 (0.504 to 0.510) | 0.510 (0.507 to 0.513) |
| AgeMen + Age1st + N_Rels | 0.505 (0.501 to 0.508) | 0.518 (0.516 to 0.521) |
| AgeMen + Age1st + N_Rels + N_Biop | 0.517 (0.514 to 0.520) | 0.526 (0.524 to 0.529) |
| AgeMen + N_Biop | 0.530 (0.526 to 0.533) | 0.505 (0.504 to 0.505) |
| AgeMen + N_Rels | 0.532 (0.529 to 0.536) | 0.523 (0.521 to 0.525) |
| AgeMen + N_Rels + N_Biop | 0.548 (0.544 to 0.551) | 0.527 (0.526 to 0.529) |
| Full_model* | 0.517 (0.514 to 0.520) | 0.527 (0.524 to 0.529) |
| N_Biop | 0.542 (0.539 to 0.546) | 0.504 (0.503 to 0.504) |
| N_Rels | 0.543 (0.540 to 0.547) | 0.522 (0.521 to 0.524) |
| N_Rels + N_Biop | 0.559 (0.555 to 0.562) | 0.526 (0.524 to 0.528) |
| **Asian, aged 30 to 49 years** |  |  |
| Age1st | 0.543 (0.530 to 0.555) |  |
| Age1st + N_Biop | 0.543 (0.530 to 0.555) |  |
| Age1st + N_Rels | 0.524 (0.511 to 0.536) | 0.501 (0.499 to 0.503) |
| Age1st + N_Rels + N_Biop | 0.524 (0.511 to 0.536) | 0.501 (0.499 to 0.503) |
| AgeMen | 0.536 (0.524 to 0.548) |  |
| AgeMen + Age1st | 0.542 (0.530 to 0.554) |  |
| AgeMen + Age1st + N_Biop | 0.542 (0.530 to 0.554) |  |
| AgeMen + Age1st + N_Rels | 0.523 (0.511 to 0.535) | 0.501 (0.499 to 0.503) |
| AgeMen + Age1st + N_Rels + N_Biop | 0.523 (0.511 to 0.535) | 0.501 (0.499 to 0.503) |
| AgeMen + N_Biop | 0.536 (0.524 to 0.548) |  |
| AgeMen + N_Rels | 0.514 (0.502 to 0.526) |  |
| AgeMen + N_Rels + N_Biop | 0.514 (0.502 to 0.526) |  |
| Full_model+ | 0.523 (0.511 to 0.535) | 0.501 (0.499 to 0.503) |
| N_Biop | 0.535 (0.522 to 0.547) |  |
| N_Rels | 0.486 (0.473 to 0.498) |  |
| N_Rels + N_Biop | 0.486 (0.473 to 0.498) |  |
| **Asian, aged 50 to 80 years** |  |  |
| Age1st | 0.537 (0.525 to 0.549) |  |
| Age1st + N_Biop | 0.537 (0.525 to 0.549) |  |
| Age1st + N_Rels | 0.556 (0.544 to 0.568) | 0.510 (0.507 to 0.514) |
| Age1st + N_Rels + N_Biop | 0.556 (0.544 to 0.568) | 0.510 (0.507 to 0.514) |
| AgeMen | 0.510 (0.498 to 0.522) |  |
| AgeMen + Age1st | 0.536 (0.524 to 0.548) |  |
| AgeMen + Age1st + N_Biop | 0.536 (0.524 to 0.548) |  |
| AgeMen + Age1st + N_Rels | 0.554 (0.543 to 0.566) | 0.512 (0.507 to 0.516) |
| AgeMen + Age1st + N_Rels + N_Biop | 0.554 (0.543 to 0.566) | 0.512 (0.507 to 0.516) |
| AgeMen + N_Biop | 0.510 (0.498 to 0.522) |  |
| AgeMen + N_Rels | 0.531 (0.519 to 0.543) |  |
| AgeMen + N_Rels + N_Biop | 0.531 (0.519 to 0.543) |  |
| Full_model* | 0.554 (0.543 to 0.566) | 0.512 (0.507 to 0.516) |
| N_Biop | 0.512 (0.500 to 0.523) |  |
| N_Rels | 0.534 (0.522 to 0.545) |  |
| N_Rels + N_Biop | 0.534 (0.522 to 0.545) |  |

### **Supplementary Figures in Main Text**

#### Supplementary Figure 1. Flowchart of individuals selected.

| **Individuals with valid genotype information** | |  |  |
| --- | --- | --- | --- |
| n=246,288 | |  |  |
|  |  |  | **Exclude, n=65,890** |
|  |  |  | Missing age at interview (age) for controls and age at diagnosis (age_dx) for cases, n=5,566 Unknown invasiveness status of breast cancer diagnosis, n=2,103  Age<30, n=4,089 Age>80, n=2,783 Not Asian or European, n=9,893 Studies with a missing value rate of 50% or higher for 2 out of 3 for variables: age of menarche, age at first live birth, and family history of breast cancer, n=41,456 |
| **Analytical dataset** | |  |  |
| n=180,398 | |  |  |

#### Supplementary Figure 2. Distribution of polygenic risk scores (scoresum) and 5-year absolute risk by country, for European ancestry.

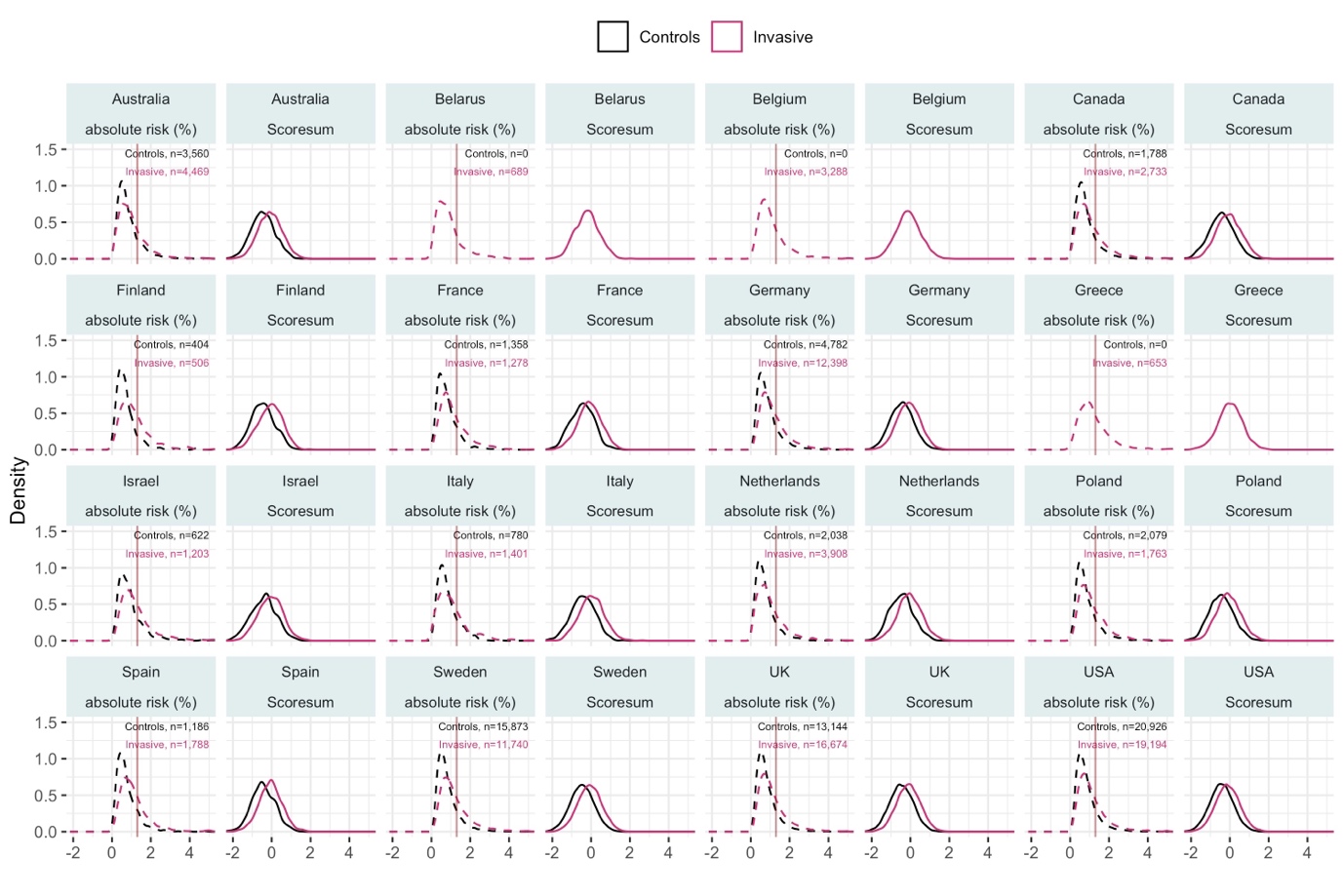

#### Supplementary Figure 3. Distribution of polygenic risk scores (scoresum) and 5-year absolute risk by country, for Asian ancestry.

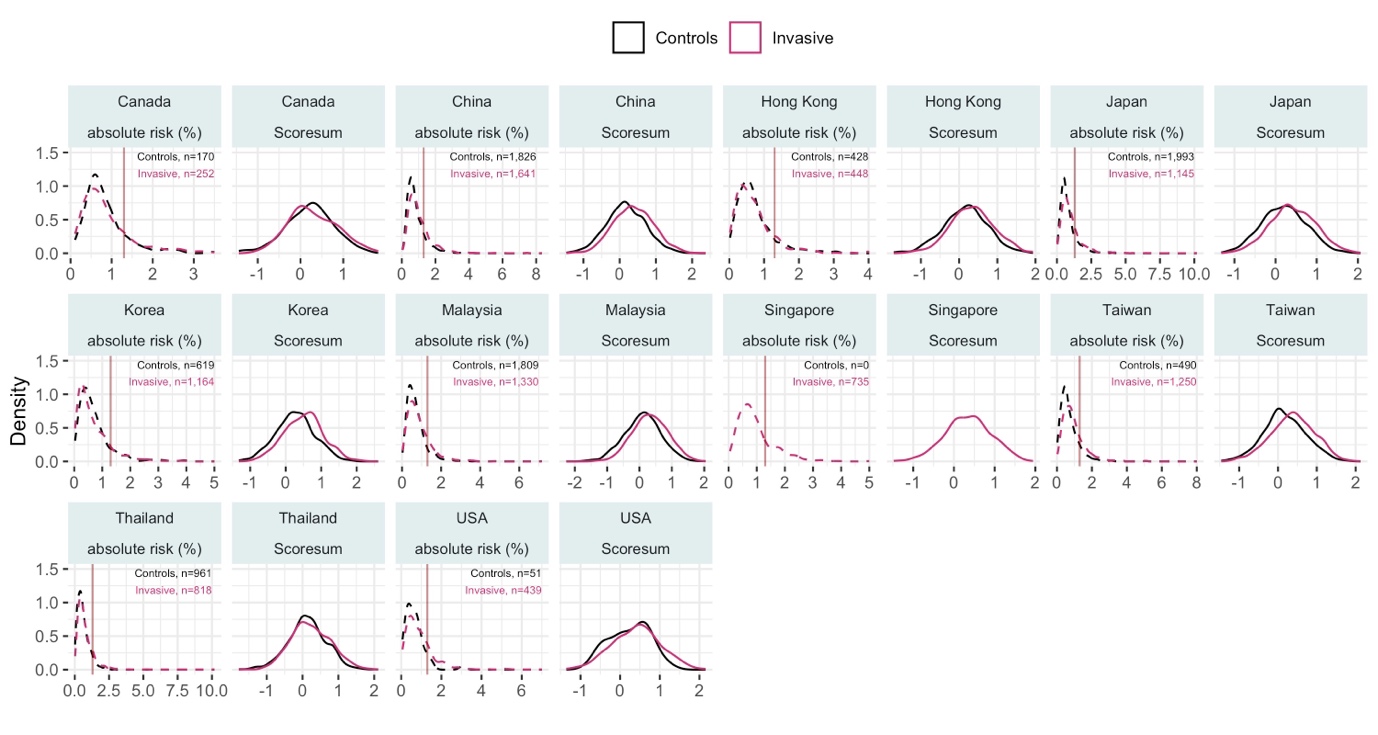

#### Supplementary Figure 4. Discriminatory ability of combinations of risk factors in the Gail model. Individuals with 5-year absolute risk ≥1.66% were identified as high risk. Five-year absolute risk was calculated using the R package “BCRA” and used to predict the case-control status of the individuals. Dots represents risk factors included in the model, and crosses indicate the model with all risk factors with the addition of atypical hyperplasia.

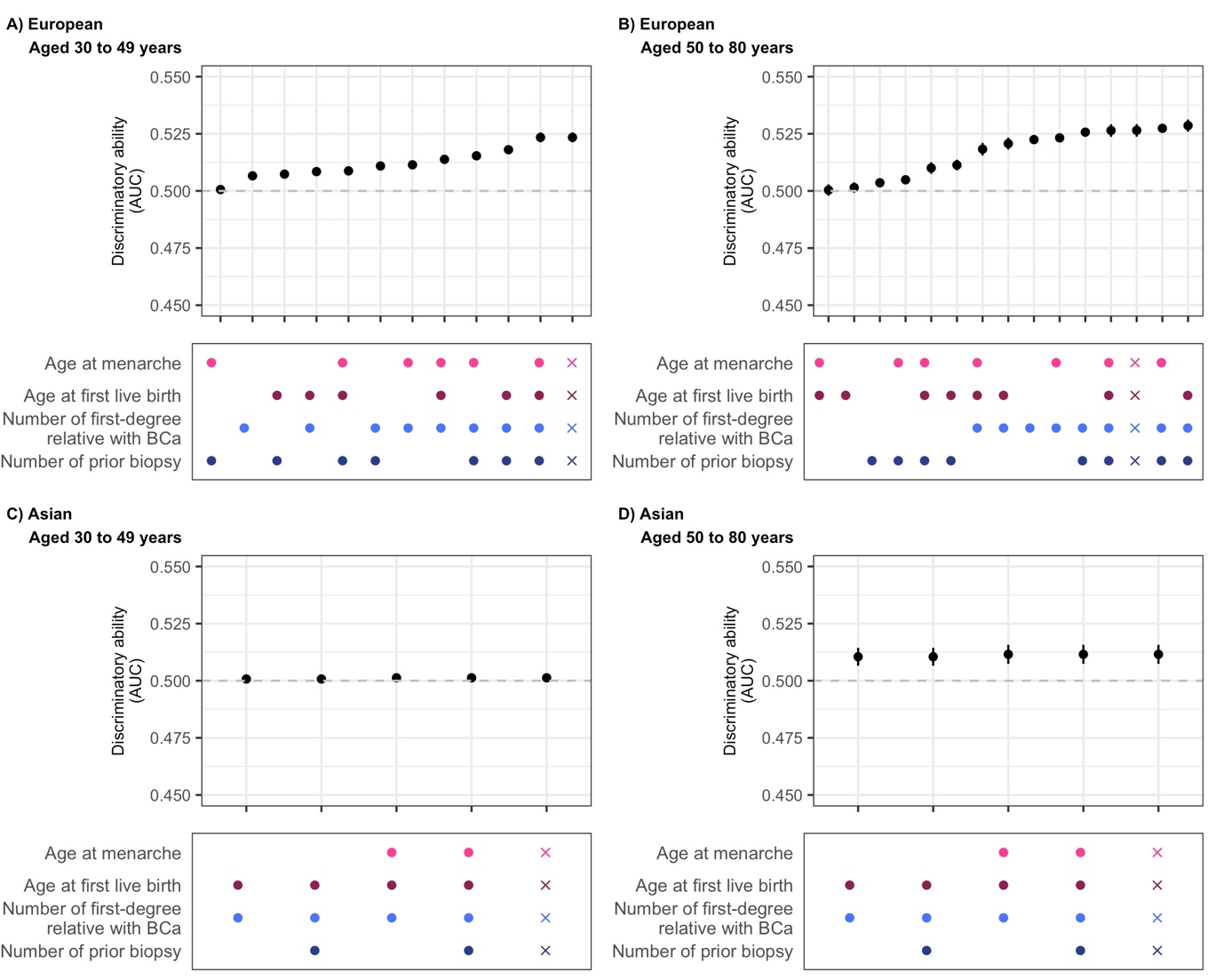

### **Additional Materials**

#### Additional Materials - Methods

##### Criteria to identify women at high risk of breast cancer

Four criteria were used to identify women at high risk of breast cancer: 1) 5-year absolute risk ≥1.66% by the Gail model [GAIL], 2) family history for breast cancer [FH] , 3) 5-year absolute risk ≥1.66% by a 313-variant breast cancer polygenic risk score [PRS], and in a subset of women 4) carriers of pathogenic variants in breast cancer predisposition genes [PTV].

##### Non-genetic breast cancer risk factors

Due to the large number of studies with varying degrees of missing data for different risk factors, the parsimonious Gail model where most studies would have information on was selected [1]. The Gail model uses information on reproductive risk factors (age of menarche, age at first live birth), personal history (number of breast biopsies, and history of atypical hyperplasia) and family (first-degree) history of breast cancer. The R package “BCRA” (version 2.1.2) was used to calculate 5-year absolute risk [1]. In addition, family history (yes/no) was separately studied. In our analysis, those with unknown family history were considered to have no family history.

##### Genetic breast cancer risk factors

We studied genetic risk based on common germline variants associated with breast cancer, using the breast cancer PRS with 313 variants calculated with plink (version 3) with the scoresum option [2, 3]. The 5-year absolute risk was obtained by applying the breast cancer incidence rates and mortality rates of “Whites” and “Chinese”, from the R package “BCRA”, to the European and Asian genetic subgroups, respectively. The rates from the “BCRA” package were used to maintain comparability between the absolute risk calculated for the PRS and the Gail model. Details to calculate absolute risk are published by Mavaddat et al. [3]. In brief, an individual's PRS percentile was obtained from the standardised PRS using the R package “pnorm”. Standardisation was done using the ancestry-specific means and standard deviations of the controls (**Additional Material - Supplementary Table 1**). The 5-year absolute risk is calculated by estimating the theoretical odds ratio of this percentile in relation to the 40-60 percentile, which is taken to represent the general population [4].

A subgroup of individuals (n_European_=56,387, n_Asian_=3,617) had both genotyping and targeted-sequencing data. Nine breast cancer predisposition genes (PTVs in *ATM, BRCA1, BRCA2, CHEK2, PALB2, BARD1, RAD51C, RAD51D,* or *TP53*) were studied collectively. PTVs disrupt the translation of these important genes, caused by three different mutations: nonsense single-nucleotide variants (SNVs), frameshift insertions or deletions (indels), and splice-disrupting SNVs. To avoid including variants that do not lead to nonsense-mediated decay, we excluded PTVs occurring in the last exon of each gene. The Centre for Cancer Genetic Epidemiology University of Cambridge) managed the preparation of the germline DNA in target-enriched sequencing libraries [5]. Information on the library preparation, sequencing, quality control, variant calling and variant annotation procedures are elaborated in Dorling et al. [5].

#### Additional Materials – Results

##### Excluded individuals

**Additional Material - Supplementary Table 2** describes the characteristics of included and excluded participants by ancestry. Excluded Europeans were younger at interview or diagnosis (included: 57 vs excluded: 54, p<0.001) The excluded women (n=41,456) from studies with a missing value rate of 50% for two out of three Gail model variables consist of 34,565 Europeans and 6,891 Asians. All excluded individuals did not have information on previous biopsies. Excluded Asians were more likely to have no information on age at menarche (included: 1,736 vs excluded: 3,024, p<0.001), age at first full-term pregnancy (included: 2,532 vs excluded: 5,465, p<0.001), and family history (included: 1,564 vs excluded: 5,803, p<0.001).

In addition to age at first full-term pregnancy (included: 24,248 vs excluded: 29,604, p<0.001), excluded Europeans were more likely to have no information on age at menarche (included: 24,107 vs excluded: 26,132, p<0.001). The 5-year absolute risk by the Gail model was significantly lower for excluded individuals across all subgroups (p<0.001).

The sumscore PRS distribution was significantly higher for excluded Europeans (included: -0.247 vs excluded: -0.237, p=0.002) and excluded Asians (included: 0.279 vs excluded: 0.308, p=0.002). The 5-year absolute risk for PRS is significantly higher for excluded Asians (excluded: 0.716 vs included: 0.681, p<0.001).

**Additional Material - Supplementary Tables 3 to 6** describe the characteristics of included and excluded participants further stratified by age (younger, 30 to 49 years; older, 50 to 80 years) and disease status (non-breast cancer controls, invasive breast cancer, DCIS). The PRS distribution (sumscore or 5-year absolute risk) was significantly higher in excluded older European invasive breast cancer patients than those included (included: -0.107 vs excluded: -0.083, p<0.001). Younger European controls and invasive breast cancer patients had significantly lower 5-year absolute risk by PRS. Apart from DCIS patients, excluded Asians had higher PRS (sumscore or 5-year absolute risk) than included individuals (p<0.05).

##### Additional Material - Supplementary Table 1. Mean and standard deviation of polygenic risk score in non-breast cancer controls from Europeans and Asians, before exclusions of studies with high missing rates for Gail model risk factors.

| **Region/ country** | **n** | **Mean** | **Standard deviation** |
| --- | --- | --- | --- |
| **Region** |  |  |  |
| European | 96,726 | -0.412 | 0.621 |
| Asian | 13,580 | 0.197 | 0.556 |
| **European population** |  |  |  |
| Australia | 4,231 | -0.437 | 0.62 |
| Belarus | 344 | -0.409 | 0.676 |
| Belgium | 1,887 | -0.444 | 0.601 |
| Canada | 2,402 | -0.425 | 0.633 |
| Denmark | 5,057 | -0.468 | 0.602 |
| Finland | 865 | -0.452 | 0.61 |
| France | 1,543 | -0.424 | 0.621 |
| Germany | 7,123 | -0.432 | 0.615 |
| Greece | 309 | -0.292 | 0.607 |
| Ireland | 86 | -0.446 | 0.611 |
| Israel | 719 | -0.377 | 0.644 |
| Italy | 1,723 | -0.34 | 0.619 |
| Netherlands | 2,600 | -0.4 | 0.631 |
| Norway | 258 | -0.368 | 0.595 |
| Poland | 2,684 | -0.424 | 0.628 |
| Republic of North Macedonia | 88 | -0.28 | 0.591 |
| Russia | 115 | -0.336 | 0.688 |
| Spain | 2,227 | -0.386 | 0.619 |
| Sweden | 16,853 | -0.447 | 0.619 |
| UK | 20,355 | -0.401 | 0.623 |
| USA | 25,257 | -0.383 | 0.621 |
| **Asian population** |  |  |  |
| Canada | 252 | 0.291 | 0.573 |
| China | 1,871 | 0.178 | 0.544 |
| Hong Kong | 531 | 0.199 | 0.581 |
| Japan | 2,162 | 0.223 | 0.548 |
| Korea | 3,201 | 0.275 | 0.551 |
| Malaysia | 1,858 | 0.092 | 0.566 |
| Singapore | 932 | 0.152 | 0.554 |
| Taiwan | 641 | 0.232 | 0.555 |
| Thailand | 1,080 | 0.114 | 0.537 |
| USA | 1,051 | 0.212 | 0.553 |

##### Additional Material - Supplementary Table 2. Demographic and risk factors differences between included and excluded individuals.

|  | **European** | | | |  | **Asian** | | | |
| --- | --- | --- | --- | --- | --- | --- | --- | --- | --- |
|  | **Included and excluded, n=196,414** | **Included, n=161,849 (82%)** | **Excluded, n=34,565 (18%)** |  |  | **Included and excluded, n=25,440** | **Included, n=18,549 (73%)** | **Excluded, n=6,891 (27%)** |  |
| **Median age (at interview/ diagnosis), years (IQR)** | 56 (48 to 64) | 57 (49 to 65) | 54 (45 to 63) | <0.001 |  | 50 (44 to 58) | 50 (44 to 58) | 50 (44 to 57) | 0.062 |
| **Age at menarche, n (%)** |  |  |  |  |  |  |  |  |  |
| <12 years | 23,742 (12) | 22,417 (14) | 1,325 (4) | <0.001 |  | 1,543 (6) | 1,155 (6) | 388 (6) | <0.001 |
| 12 to 14 years | 72,071 (37) | 68,265 (42) | 3,806 (11) |  |  | 8,159 (32) | 6,684 (36) | 1,475 (21) |  |
| >=14 years | 50,362 (26) | 47,060 (29) | 3,302 (10) |  |  | 10,978 (43) | 8,974 (48) | 2,004 (29) |  |
| Unknown | 50,239 (26) | 24,107 (15) | 26,132 (76) |  |  | 4,760 (19) | 1,736 (9) | 3,024 (44) |  |
| **Age at first full term pregnancy, n (%)** |  |  |  |  |  |  |  |  |  |
| Nulliparous | 22,988 (12) | 20,608 (13) | 2,380 (7) | <0.001 |  | 2,850 (11) | 2,399 (13) | 451 (7) | <0.001 |
| <20 years | 12,620 (6) | 12,359 (8) | 261 (1) |  |  | 736 (3) | 707 (4) | 29 (0) |  |
| 20 to 24 years | 47,710 (24) | 46,689 (29) | 1,021 (3) |  |  | 4,267 (17) | 3,893 (21) | 374 (5) |  |
| 25 to 29 years | 38,960 (20) | 38,099 (24) | 861 (2) |  |  | 6,635 (26) | 6,159 (33) | 476 (7) |  |
| >=30 years | 20,284 (10) | 19,846 (12) | 438 (1) |  |  | 2,955 (12) | 2,859 (15) | 96 (1) |  |
| Unknown | 53,852 (27) | 24,248 (15) | 29,604 (86) |  |  | 7,997 (31) | 2,532 (14) | 5,465 (79) |  |
| **Number of first degree family history of breast cancer, n (%)** | | | | | | | | | |
| No | 106,831 (54) | 98,537 (61) | 8,294 (24) | <0.001 |  | 16,449 (65) | 15,389 (83) | 1,060 (15) | <0.001 |
| 1 | 19,065 (10) | 16,719 (10) | 2,346 (7) |  |  | 1,455 (6) | 1,427 (8) | 28 (0) |  |
| 2+ | 3,447 (2) | 3,160 (2) | 287 (1) |  |  | 169 (1) | 169 (1) | 0 (0) |  |
| Unknown | 67,071 (34) | 43,433 (27) | 23,638 (68) |  |  | 7,367 (29) | 1,564 (8) | 5,803 (84) |  |
| **Number of breast biopsy, n (%)** |  |  |  |  |  |  |  |  |  |
| No | 4,207 (2) | 4,207 (3) | 0 (0) |  |  | 0 (0) | 0 (0) | 0 (0) |  |
| 1 | 3,641 (2) | 3,641 (2) | 0 (0) |  |  | 0 (0) | 0 (0) | 0 (0) |  |
| 2+ | 2,073 (1) | 2,073 (1) | 0 (0) |  |  | 0 (0) | 0 (0) | 0 (0) |  |
| Unknown | 186,493 (95) | 151,928 (94) | 34,565 (100) |  |  | 25,440 (100) | 18,549 (100) | 6,891 (100) |  |
| **Atypical hyperplasia, n (%)** |  |  |  |  |  |  |  |  |  |
| No | 4,207 (2) | 4,207 (3) | 0 (0) |  |  | 0 (0) | 0 (0) | 0 (0) |  |
| Yes | 62 (0) | 62 (0) | 0 (0) |  |  | 0 (0) | 0 (0) | 0 (0) |  |
| Unknown | 192,145 (98) | 157,580 (97) | 34,565 (100) |  |  | 25,440 (100) | 18,549 (100) | 6,891 (100) |  |
| **Median 5-year absolute risk by the Gail model (IQR)** | 1.154 (0.833 to 1.597) | 1.248 (0.910 to 1.699) | 0.842 (0.575 to 1.120) | <0.001 |  | 0.513 (0.382 to 0.730) | 0.609 (0.447 to 0.793) | 0.413 (0.350 to 0.477) | <0.001 |
| **Protein truncating variants (9 Genes)** |  |  |  |  |  |  |  |  |  |
| No | 61,433 (31) | 53,803 (33) | 7,630 (22) | 0.538 |  | 3,457 (14) | 3,457 (19) | 0 (0) |  |
| Yes | 2,937 (1) | 2,584 (2) | 353 (1) |  |  | 160 (1) | 160 (1) | 0 (0) |  |
| Unknown | 132,044 (67) | 105,462 (65) | 26,582 (77) |  |  | 21,823 (86) | 14,932 (81) | 6,891 (100) |  |
| **Polygenic risk score (PRS)** | -0.245 (-0.676 to 0.189) | -0.247 (-0.677 to 0.186) | -0.237 (-0.673 to 0.201) | 0.002 |  | 0.287 (-0.087 to 0.676) | 0.279 (-0.097 to 0.675) | 0.308 (-0.056 to 0.678) | 0.002 |
| **Median 5-year absolute risk by PRS (IQR)** | 0.821 (0.525 to 1.264) | 0.829 (0.536 to 1.273) | 0.777 (0.479 to 1.223) | <0.001 |  | 0.691 (0.430 to 1.069) | 0.681 (0.419 to 1.063) | 0.716 (0.463 to 1.088) | <0.001 |

##### Additional Material - Supplementary Table 3. Demographic and risk factors differences between included (Yes) and excluded (No) individuals, in European aged 30 to 49 years.

|  | **Non-breast cancer controls** | | | |  | **Invasive breast cancer cases** | | | |  | **DCIS** | | | |
| --- | --- | --- | --- | --- | --- | --- | --- | --- | --- | --- | --- | --- | --- | --- |
|  | All, n=22,346 | Yes, n=17,082 | No, n=5,264 |  |  | All, n=30,090 | Yes, n=23,224 | No, n=6,866 |  |  | All, n=2,826 | Yes, n=2,342 | No, n=484 |  |
| **Median age (at interview/ diagnosis), years (IQR)** | 44 (40 to 47) | 44 (41 to 47) | 42 (37 to 46) | <0.001 |  | 44 (39 to 47) | 44 (40 to 47) | 42 (38 to 46) | <0.001 |  | 45 (41 to 47) | 45 (42 to 47) | 44 (40 to 47) | 0.003 |
| **Age at menarche, n (%)** |  |  |  |  |  |  |  |  |  |  |  |  |  |  |
| <12 | 2,655 (12) | 2,531 (15) | 124 (2) | 0.636 |  | 3,416 (11) | 3,150 (14) | 266 (4) | <0.001 |  | 472 (17) | 404 (17) | 68 (14) | <0.001 |
| 12 to 14 | 8,089 (36) | 7,731 (45) | 358 (7) |  |  | 10,359 (34) | 9,603 (41) | 756 (11) |  |  | 1,199 (42) | 1,105 (47) | 94 (19) |  |
| >=14 | 4,711 (21) | 4,486 (26) | 225 (4) |  |  | 5,854 (19) | 5,328 (23) | 526 (8) |  |  | 688 (24) | 631 (27) | 57 (12) |  |
| Unknown | 6,891 (31) | 2,334 (14) | 4,557 (87) |  |  | 10,461 (35) | 5,143 (22) | 5,318 (77) |  |  | 467 (17) | 202 (9) | 265 (55) |  |
| **Age at first full term pregnancy, n (%)** |  |  |  |  |  |  |  |  |  |  |  |  |  |  |
| Nulliparous | 3,171 (14) | 2,588 (15) | 583 (11) | <0.001 |  | 4,038 (13) | 3,737 (16) | 301 (4) | <0.001 |  | 495 (18) | 452 (19) | 43 (9) | <0.001 |
| <20 | 1,052 (5) | 1,031 (6) | 21 (0) |  |  | 1,351 (4) | 1,322 (6) | 29 (0) |  |  | 122 (4) | 122 (5) | 0 (0) |  |
| 20 to 24 | 3,953 (18) | 3,866 (23) | 87 (2) |  |  | 4,369 (15) | 4,285 (18) | 84 (1) |  |  | 494 (17) | 491 (21) | 3 (1) |  |
| 25 to 29 | 4,545 (20) | 4,432 (26) | 113 (2) |  |  | 5,145 (17) | 5,008 (22) | 137 (2) |  |  | 595 (21) | 590 (25) | 5 (1) |  |
| >=30 | 3,145 (14) | 3,083 (18) | 62 (1) |  |  | 4,031 (13) | 3,902 (17) | 129 (2) |  |  | 481 (17) | 476 (20) | 5 (1) |  |
| Unknown | 6,480 (29) | 2,082 (12) | 4,398 (84) |  |  | 11,156 (37) | 4,970 (21) | 6,186 (90) |  |  | 639 (23) | 211 (9) | 428 (88) |  |
| **Number of first degree family history of breast cancer, n (%)** |  |  |  |  |  |  |  |  |  |  |  |  |  |  |
| No | 12,697 (57) | 11,340 (66) | 1,357 (26) | 0.078 |  | 16,162 (54) | 14,098 (61) | 2,064 (30) | <0.001 |  | 966 (34) | 905 (39) | 61 (13) | 0.002 |
| 1 | 1,430 (6) | 1,295 (8) | 135 (3) |  |  | 4,140 (14) | 3,370 (15) | 770 (11) |  |  | 395 (14) | 348 (15) | 47 (10) |  |
| 2+ | 146 (1) | 137 (1) | 9 (0) |  |  | 623 (2) | 537 (2) | 86 (1) |  |  | 90 (3) | 80 (3) | 10 (2) |  |
| Unknown | 8,073 (36) | 4,310 (25) | 3,763 (71) |  |  | 9,165 (30) | 5,219 (22) | 3,946 (57) |  |  | 1,375 (49) | 1,009 (43) | 366 (76) |  |
| **Number of breast biopsy, n (%)** |  |  |  |  |  |  |  |  |  |  |  |  |  |  |
| No | 301 (1) | 301 (2) | 0 (0) |  |  | 1,310 (4) | 1,310 (6) | 0 (0) |  |  | 43 (2) | 43 (2) | 0 (0) |  |
| 1 | 74 (0) | 74 (0) | 0 (0) |  |  | 784 (3) | 784 (3) | 0 (0) |  |  | 60 (2) | 60 (3) | 0 (0) |  |
| 2+ | 23 (0) | 23 (0) | 0 (0) |  |  | 532 (2) | 532 (2) | 0 (0) |  |  | 42 (1) | 42 (2) | 0 (0) |  |
| Unknown | 21,948 (98) | 16,684 (98) | 5,264 (100) |  |  | 27,464 (91) | 20,598 (89) | 6,866 (100) |  |  | 2,681 (95) | 2,197 (94) | 484 (100) |  |
| **Atypical hyperplasia, n (%)** |  |  |  |  |  |  |  |  |  |  |  |  |  |  |
| No | 301 (1) | 301 (2) | 0 (0) |  |  | 1,310 (4) | 1,310 (6) | 0 (0) |  |  | 43 (2) | 43 (2) | 0 (0) |  |
| Yes | 1 (0) | 1 (0) | 0 (0) |  |  | 13 (0) | 13 (0) | 0 (0) |  |  | 1 (0) | 1 (0) | 0 (0) |  |
| Unknown | 22,044 (99) | 16,780 (98) | 5,264 (100) |  |  | 28,767 (96) | 21,901 (94) | 6,866 (100) |  |  | 2,782 (98) | 2,298 (98) | 484 (100) |  |
| **Median 5-year absolute risk by the Gail model (IQR)** | 0.662 (0.467 to 0.910) | 0.746 (0.555 to 0.948) | 0.467 (0.260 to 0.587) | <0.001 |  | 0.638 (0.467 to 0.976) | 0.739 (0.522 to 1.035) | 0.502 (0.324 to 0.613) | <0.001 |  | 0.811 (0.594 to 1.059) | 0.860 (0.652 to 1.126) | 0.562 (0.433 to 0.652) | <0.001 |
| **Protein truncating variants (9 Genes)** |  |  |  |  |  |  |  |  |  |  |  |  |  |  |
| No | 5,716 (26) | 4,687 (27) | 1,029 (20) | <0.001 |  | 8,949 (30) | 7,748 (33) | 1,201 (17) | 0.687 |  | 446 (16) | 406 (17) | 40 (8) | 0.055 |
| Yes | 186 (1) | 173 (1) | 13 (0) |  |  | 939 (3) | 808 (3) | 131 (2) |  |  | 27 (1) | 21 (1) | 6 (1) |  |
| Unknown | 16,444 (74) | 12,222 (72) | 4,222 (80) |  |  | 20,202 (67) | 14,668 (63) | 5,534 (81) |  |  | 2,353 (83) | 1,915 (82) | 438 (90) |  |
| **Polygenic risk score (PRS)** | -0.429 (-0.841 to -0.016) | -0.426 (-0.842 to -0.010) | -0.436 (-0.841 to -0.033) | 0.437 |  | -0.052 (-0.479 to 0.368) | -0.058 (-0.484 to 0.367) | -0.036 (-0.459 to 0.371) | 0.066 |  | -0.072 (-0.484 to 0.334) | -0.077 (-0.484 to 0.328) | -0.053 (-0.485 to 0.376) | 0.335 |
| **Median 5-year absolute risk by PRS (IQR)** | 0.480 (0.296 to 0.748) | 0.499 (0.314 to 0.769) | 0.416 (0.238 to 0.678) | <0.001 |  | 0.668 (0.404 to 1.050) | 0.683 (0.415 to 1.068) | 0.620 (0.368 to 1.000) | <0.001 |  | 0.717 (0.469 to 1.065) | 0.718 (0.477 to 1.068) | 0.712 (0.451 to 1.063) | 0.207 |

##### Additional Material - Supplementary Table 4. Demographic and risk factors differences between included (Yes) and excluded (No) individuals, in European aged 50 to 80 years.

|  | **Non-breast cancer controls** | | | |  | **Invasive breast cancer cases** | | | |  | **DCIS** | | | |
| --- | --- | --- | --- | --- | --- | --- | --- | --- | --- | --- | --- | --- | --- | --- |
|  | All, n=61,028 | Yes, n=51,458 | No, n=9,570 |  |  | All, n=71,779 | Yes, n=60,461 | No, n=11,318 |  |  | All, n=8,345 | Yes, n=7,282 | No, n=1,063 |  |
| **Median age (at interview/ diagnosis), years (IQR)** | 61 (55 to 67) | 61 (55 to 66) | 61 (55 to 68) | <0.001 |  | 61 (55 to 67) | 61 (56 to 67) | 60 (54 to 67) | <0.001 |  | 58 (54 to 65) | 59 (54 to 65) | 57 (52 to 63) | <0.001 |
| **Age at menarche, n (%)** |  |  |  |  |  |  |  |  |  |  |  |  |  |  |
| <12 | 7,594 (12) | 7,294 (14) | 300 (3) | <0.001 |  | 8,146 (11) | 7,709 (13) | 437 (4) | <0.001 |  | 1,459 (17) | 1,329 (18) | 130 (12) | 0.439 |
| 12 to 14 | 23,307 (38) | 22,512 (44) | 795 (8) |  |  | 25,378 (35) | 23,873 (39) | 1,505 (13) |  |  | 3,739 (45) | 3,441 (47) | 298 (28) |  |
| >=14 | 17,434 (29) | 16,647 (32) | 787 (8) |  |  | 19,368 (27) | 17,841 (30) | 1,527 (13) |  |  | 2,307 (28) | 2,127 (29) | 180 (17) |  |
| Unknown | 12,693 (21) | 5,005 (10) | 7,688 (80) |  |  | 18,887 (26) | 11,038 (18) | 7,849 (69) |  |  | 840 (10) | 385 (5) | 455 (43) |  |
| **Age at first full term pregnancy, n (%)** |  |  |  |  |  |  |  |  |  |  |  |  |  |  |
| Nulliparous | 6,536 (11) | 5,731 (11) | 805 (8) | <0.001 |  | 7,558 (11) | 7,032 (12) | 526 (5) | <0.001 |  | 1,190 (14) | 1,068 (15) | 122 (11) | <0.001 |
| <20 | 4,453 (7) | 4,396 (9) | 57 (1) |  |  | 4,968 (7) | 4,816 (8) | 152 (1) |  |  | 674 (8) | 672 (9) | 2 (0) |  |
| 20 to 24 | 18,265 (30) | 17,997 (35) | 268 (3) |  |  | 18,199 (25) | 17,642 (29) | 557 (5) |  |  | 2,430 (29) | 2,408 (33) | 22 (2) |  |
| 25 to 29 | 13,306 (22) | 13,111 (25) | 195 (2) |  |  | 13,553 (19) | 13,166 (22) | 387 (3) |  |  | 1,816 (22) | 1,792 (25) | 24 (2) |  |
| >=30 | 5,471 (9) | 5,404 (11) | 67 (1) |  |  | 6,241 (9) | 6,071 (10) | 170 (2) |  |  | 915 (11) | 910 (12) | 5 (0) |  |
| Unknown | 12,997 (21) | 4,819 (9) | 8,178 (85) |  |  | 21,260 (30) | 11,734 (19) | 9,526 (84) |  |  | 1,320 (16) | 432 (6) | 888 (84) |  |
| **Number of first degree family history of breast cancer, n (%)** |  |  |  |  |  |  |  |  |  |  |  |  |  |  |
| No | 35,512 (58) | 34,289 (67) | 1,223 (13) | <0.001 |  | 38,262 (53) | 34,805 (58) | 3,457 (31) | <0.001 |  | 3,232 (39) | 3,100 (43) | 132 (12) | 0.462 |
| 1 | 4,151 (7) | 4,053 (8) | 98 (1) |  |  | 8,142 (11) | 6,886 (11) | 1,256 (11) |  |  | 807 (10) | 767 (11) | 40 (4) |  |
| 2+ | 672 (1) | 654 (1) | 18 (0) |  |  | 1,679 (2) | 1,527 (3) | 152 (1) |  |  | 237 (3) | 225 (3) | 12 (1) |  |
| Unknown | 20,693 (34) | 12,462 (24) | 8,231 (86) |  |  | 23,696 (33) | 17,243 (29) | 6,453 (57) |  |  | 4,069 (49) | 3,190 (44) | 879 (83) |  |
| **Number of breast biopsy, n (%)** |  |  |  |  |  |  |  |  |  |  |  |  |  |  |
| No | 629 (1) | 629 (1) | 0 (0) |  |  | 1,871 (3) | 1,871 (3) | 0 (0) |  |  | 53 (1) | 53 (1) | 0 (0) |  |
| 1 | 203 (0) | 203 (0) | 0 (0) |  |  | 2,364 (3) | 2,364 (4) | 0 (0) |  |  | 156 (2) | 156 (2) | 0 (0) |  |
| 2+ | 80 (0) | 80 (0) | 0 (0) |  |  | 1,290 (2) | 1,290 (2) | 0 (0) |  |  | 106 (1) | 106 (1) | 0 (0) |  |
| Unknown | 60,116 (99) | 50,546 (98) | 9,570 (100) |  |  | 66,254 (92) | 54,936 (91) | 11,318 (100) |  |  | 8,030 (96) | 6,967 (96) | 1,063 (100) |  |
| **Atypical hyperplasia, n (%)** |  |  |  |  |  |  |  |  |  |  |  |  |  |  |
| No | 629 (1) | 629 (1) | 0 (0) |  |  | 1,871 (3) | 1,871 (3) | 0 (0) |  |  | 53 (1) | 53 (1) | 0 (0) |  |
| Yes | 4 (0) | 4 (0) | 0 (0) |  |  | 36 (0) | 36 (0) | 0 (0) |  |  | 7 (0) | 7 (0) | 0 (0) |  |
| Unknown | 60,395 (99) | 50,825 (99) | 9,570 (100) |  |  | 69,872 (97) | 58,554 (97) | 11,318 (100) |  |  | 8,285 (99) | 7,222 (99) | 1,063 (100) |  |
| **Median 5-year absolute risk by the Gail model (IQR)** | 1.328 (1.065 to 1.700) | 1.404 (1.128 to 1.749) | 1.032 (0.808 to 1.147) | <0.001 |  | 1.367 (1.060 to 1.807) | 1.428 (1.109 to 1.883) | 1.060 (0.842 to 1.161) | <0.001 |  | 1.362 (1.088 to 1.749) | 1.411 (1.149 to 1.811) | 0.998 (0.773 to 1.145) | <0.001 |
| **Protein truncating variants (9 Genes)** |  |  |  |  |  |  |  |  |  |  |  |  |  |  |
| No | 21,089 (35) | 19,528 (38) | 1,561 (16) | 0.381 |  | 23,759 (33) | 20,178 (33) | 3,581 (32) | 0.071 |  | 1,474 (18) | 1,256 (17) | 218 (21) | 0.425 |
| Yes | 437 (1) | 410 (1) | 27 (0) |  |  | 1,289 (2) | 1,119 (2) | 170 (2) |  |  | 59 (1) | 53 (1) | 6 (1) |  |
| Unknown | 39,502 (65) | 31,520 (61) | 7,982 (83) |  |  | 46,731 (65) | 39,164 (65) | 7,567 (67) |  |  | 6,812 (82) | 5,973 (82) | 839 (79) |  |
| **Polygenic risk score (PRS)** | -0.459 (-0.870 to -0.046) | -0.459 (-0.870 to -0.046) | -0.459 (-0.868 to -0.045) | 0.645 |  | -0.103 (-0.514 to 0.314) | -0.107 (-0.517 to 0.309) | -0.083 (-0.496 to 0.346) | <0.001 |  | -0.172 (-0.577 to 0.248) | -0.170 (-0.572 to 0.248) | -0.177 (-0.607 to 0.238) | 0.488 |
| **Median 5-year absolute risk by PRS (IQR)** | 0.763 (0.520 to 1.133) | 0.763 (0.519 to 1.130) | 0.768 (0.523 to 1.148) | 0.142 |  | 1.058 (0.720 to 1.571) | 1.055 (0.718 to 1.568) | 1.069 (0.727 to 1.585) | 0.05 |  | 0.968 (0.668 to 1.447) | 0.971 (0.670 to 1.448) | 0.945 (0.646 to 1.419) | 0.132 |

##### Additional Material - Supplementary Table 5. Demographic and risk factors differences between included (Yes) and excluded (No) individuals, in Asian aged 30 to 49 years.

|  | **Non-breast cancer controls** | | | |  | **Invasive breast cancer cases** | | | |  | **DCIS** | |
| --- | --- | --- | --- | --- | --- | --- | --- | --- | --- | --- | --- | --- |
|  | All, n=5,653 | Yes, n=3,822 | No, n=1,831 |  |  | All, n=6,173 | Yes, n=4,634 | No, n=1,539 |  |  | All, n=500 | Yes, n=500 |
| **Median age (at interview/ diagnosis), years (IQR)** | 44 (40 to 47) | 44 (40 to 47) | 44 (41 to 47) | <0.001 |  | 43 (38 to 46) | 43 (38 to 47) | 44 (40 to 46) | <0.001 |  | 43 (38 to 47) | 43 (38 to 47) |
| **Age at menarche, n (%)** |  |  |  |  |  |  |  |  |  |  |  |  |
| <12 | 424 (8) | 323 (8) | 101 (6) | 0.225 |  | 437 (7) | 377 (8) | 60 (4) | <0.001 |  | 38 (8) | 38 (8) |
| 12 to 14 | 2,027 (36) | 1,523 (40) | 504 (28) |  |  | 1,980 (32) | 1,774 (38) | 206 (13) |  |  | 185 (37) | 185 (37) |
| >=14 | 2,386 (42) | 1,747 (46) | 639 (35) |  |  | 2,216 (36) | 1,897 (41) | 319 (21) |  |  | 220 (44) | 220 (44) |
| Unknown | 816 (14) | 229 (6) | 587 (32) |  |  | 1,540 (25) | 586 (13) | 954 (62) |  |  | 57 (11) | 57 (11) |
| **Age at first full term pregnancy, n (%)** |  |  |  |  |  |  |  |  |  |  |  |  |
| Nulliparous | 752 (13) | 629 (16) | 123 (7) | <0.001 |  | 896 (15) | 791 (17) | 105 (7) | <0.001 |  | 108 (22) | 108 (22) |
| <20 | 142 (3) | 134 (4) | 8 (0) |  |  | 127 (2) | 127 (3) | 0 (0) |  |  | 13 (3) | 13 (3) |
| 20 to 24 | 844 (15) | 684 (18) | 160 (9) |  |  | 708 (11) | 707 (15) | 1 (0) |  |  | 40 (8) | 40 (8) |
| 25 to 29 | 1,725 (31) | 1,423 (37) | 302 (16) |  |  | 1,578 (26) | 1,578 (34) | 0 (0) |  |  | 142 (28) | 142 (28) |
| >=30 | 690 (12) | 625 (16) | 65 (4) |  |  | 838 (14) | 838 (18) | 0 (0) |  |  | 81 (16) | 81 (16) |
| Unknown | 1,500 (27) | 327 (9) | 1,173 (64) |  |  | 2,026 (33) | 593 (13) | 1,433 (93) |  |  | 116 (23) | 116 (23) |
| **Number of first degree family history of breast cancer, n (%)** |  |  |  |  |  |  |  |  |  |  |  |  |
| No | 3,692 (65) | 3,389 (89) | 303 (17) | 0.249 |  | 4,136 (67) | 3,858 (83) | 278 (18) | <0.001 |  | 360 (72) | 360 (72) |
| 1 | 223 (4) | 207 (5) | 16 (1) |  |  | 434 (7) | 434 (9) | 0 (0) |  |  | 63 (13) | 63 (13) |
| 2+ | 28 (0) | 28 (1) | 0 (0) |  |  | 44 (1) | 44 (1) | 0 (0) |  |  | 6 (1) | 6 (1) |
| Unknown | 1,710 (30) | 198 (5) | 1,512 (83) |  |  | 1,559 (25) | 298 (6) | 1,261 (82) |  |  | 71 (14) | 71 (14) |
| **Number of breast biopsy, n (%)** |  |  |  |  |  |  |  |  |  |  |  |  |
| No | 0 (0) | 0 (0) | 0 (0) |  |  | 0 (0) | 0 (0) | 0 (0) |  |  | 0 (0) | 0 (0) |
| 1 | 0 (0) | 0 (0) | 0 (0) |  |  | 0 (0) | 0 (0) | 0 (0) |  |  | 0 (0) | 0 (0) |
| 2+ | 0 (0) | 0 (0) | 0 (0) |  |  | 0 (0) | 0 (0) | 0 (0) |  |  | 0 (0) | 0 (0) |
| Unknown | 5,653 (100) | 3,822 (100) | 1,831 (100) |  |  | 6,173 (100) | 4,634 (100) | 1,539 (100) |  |  | 500 (100) | 500 (100) |
| **Atypical hyperplasia, n (%)** |  |  |  |  |  |  |  |  |  |  |  |  |
| No | 0 (0) | 0 (0) | 0 (0) |  |  | 0 (0) | 0 (0) | 0 (0) |  |  | 0 (0) | 0 (0) |
| Yes | 0 (0) | 0 (0) | 0 (0) |  |  | 0 (0) | 0 (0) | 0 (0) |  |  | 0 (0) | 0 (0) |
| Unknown | 5,653 (100) | 3,822 (100) | 1,831 (100) |  |  | 6,173 (100) | 4,634 (100) | 1,539 (100) |  |  | 500 (100) | 500 (100) |
| **Median 5-year absolute risk by the Gail model (IQR)** | 0.422 (0.322 to 0.610) | 0.501 (0.344 to 0.652) | 0.351 (0.296 to 0.408) | <0.001 |  | 0.368 (0.262 to 0.610) | 0.466 (0.294 to 0.654) | 0.322 (0.217 to 0.351) | <0.001 |  | 0.446 (0.269 to 0.647) | 0.446 (0.269 to 0.647) |
| **Protein truncating variants (9 Genes)** |  |  |  |  |  |  |  |  |  |  |  |  |
| No | 751 (13) | 751 (20) | 0 (0) |  |  | 1,305 (21) | 1,305 (28) | 0 (0) |  |  | 184 (37) | 184 (37) |
| Yes | 18 (0) | 18 (0) | 0 (0) |  |  | 95 (2) | 95 (2) | 0 (0) |  |  | 5 (1) | 5 (1) |
| Unknown | 4,884 (86) | 3,053 (80) | 1,831 (100) |  |  | 4,773 (77) | 3,234 (70) | 1,539 (100) |  |  | 311 (62) | 311 (62) |
| **Polygenic risk score (PRS)** | 0.203 (-0.157 to 0.559) | 0.181 (-0.169 to 0.544) | 0.234 (-0.127 to 0.595) | 0.003 |  | 0.407 (0.024 to 0.789) | 0.393 (-0.002 to 0.785) | 0.452 (0.101 to 0.806) | <0.001 |  | 0.414 (-0.008 to 0.799) | 0.414 (-0.008 to 0.799) |
| **Median 5-year absolute risk by PRS (IQR)** | 0.506 (0.316 to 0.770) | 0.489 (0.295 to 0.743) | 0.541 (0.362 to 0.818) | <0.001 |  | 0.572 (0.329 to 0.925) | 0.547 (0.311 to 0.906) | 0.638 (0.409 to 0.976) | <0.001 |  | 0.532 (0.335 to 0.899) | 0.532 (0.335 to 0.899) |

##### Additional Material - Supplementary Table 6. Demographic and risk factors differences between included (Yes) and excluded (No) individuals, in Asian aged 50 to 80 years.

|  | **Non-breast cancer controls** | | | |  | **Invasive breast cancer cases** | | | |  | **DCIS** | | | |
| --- | --- | --- | --- | --- | --- | --- | --- | --- | --- | --- | --- | --- | --- | --- |
|  | All, n=6,672 | Yes, n=4,525 | No, n=2,147 |  |  | All, n=5,957 | Yes, n=4,588 | No, n=1,369 |  |  | All, n=485 | Yes, n=480 | No, n=5 |  |
| **Median age (at interview/ diagnosis), years (IQR)** | 57 (53 to 63) | 57 (53 to 63) | 57 (53 to 63) | 0.305 |  | 57 (53 to 63) | 57 (53 to 64) | 57 (53 to 63) | 0.565 |  | 56 (52 to 62) | 56 (52 to 62) | 57 (52 to 72) | 0.718 |
| **Age at menarche, n (%)** |  |  |  |  |  |  |  |  |  |  |  |  |  |  |
| <12 | 337 (5) | 185 (4) | 152 (7) | <0.001 |  | 284 (5) | 209 (5) | 75 (5) | <0.001 |  | 23 (5) | 23 (5) | 0 (0) | 0.231 |
| 12 to 14 | 2,053 (31) | 1,557 (34) | 496 (23) |  |  | 1,770 (30) | 1,504 (33) | 266 (19) |  |  | 144 (30) | 141 (29) | 3 (60) |  |
| >=14 | 3,254 (49) | 2,574 (57) | 680 (32) |  |  | 2,654 (45) | 2,289 (50) | 365 (27) |  |  | 248 (51) | 247 (51) | 1 (20) |  |
| Unknown | 1,028 (15) | 209 (5) | 819 (38) |  |  | 1,249 (21) | 586 (13) | 663 (48) |  |  | 70 (14) | 69 (14) | 1 (20) |  |
| **Age at first full term pregnancy, n (%)** |  |  |  |  |  |  |  |  |  |  |  |  |  |  |
| Nulliparous | 491 (7) | 376 (8) | 115 (5) | <0.001 |  | 552 (9) | 445 (10) | 107 (8) | <0.001 |  | 51 (11) | 50 (10) | 1 (20) | 0.157 |
| <20 | 203 (3) | 182 (4) | 21 (1) |  |  | 235 (4) | 235 (5) | 0 (0) |  |  | 16 (3) | 16 (3) | 0 (0) |  |
| 20 to 24 | 1,477 (22) | 1,264 (28) | 213 (10) |  |  | 1,103 (19) | 1,103 (24) | 0 (0) |  |  | 95 (20) | 95 (20) | 0 (0) |  |
| 25 to 29 | 1,587 (24) | 1,413 (31) | 174 (8) |  |  | 1,452 (24) | 1,452 (32) | 0 (0) |  |  | 151 (31) | 151 (31) | 0 (0) |  |
| >=30 | 555 (8) | 524 (12) | 31 (1) |  |  | 716 (12) | 716 (16) | 0 (0) |  |  | 75 (15) | 75 (16) | 0 (0) |  |
| Unknown | 2,359 (35) | 766 (17) | 1,593 (74) |  |  | 1,899 (32) | 637 (14) | 1,262 (92) |  |  | 97 (20) | 93 (19) | 4 (80) |  |
| **Number of first degree family history of breast cancer, n (%)** |  |  |  |  |  |  |  |  |  |  |  |  |  |  |
| No | 4,077 (61) | 3,792 (84) | 285 (13) | 0.131 |  | 3,836 (64) | 3,642 (79) | 194 (14) | <0.001 |  | 348 (72) | 348 (72) | 0 (0) |  |
| 1 | 242 (4) | 230 (5) | 12 (1) |  |  | 442 (7) | 442 (10) | 0 (0) |  |  | 51 (11) | 51 (11) | 0 (0) |  |
| 2+ | 35 (1) | 35 (1) | 0 (0) |  |  | 49 (1) | 49 (1) | 0 (0) |  |  | 7 (1) | 7 (1) | 0 (0) |  |
| Unknown | 2,318 (35) | 468 (10) | 1,850 (86) |  |  | 1,630 (27) | 455 (10) | 1,175 (86) |  |  | 79 (16) | 74 (15) | 5 (100) |  |
| **Number of breast biopsy, n (%)** |  |  |  |  |  |  |  |  |  |  |  |  |  |  |
| No | 0 (0) | 0 (0) | 0 (0) |  |  | 0 (0) | 0 (0) | 0 (0) |  |  | 0 (0) | 0 (0) | 0 (0) |  |
| 1 | 0 (0) | 0 (0) | 0 (0) |  |  | 0 (0) | 0 (0) | 0 (0) |  |  | 0 (0) | 0 (0) | 0 (0) |  |
| 2+ | 0 (0) | 0 (0) | 0 (0) |  |  | 0 (0) | 0 (0) | 0 (0) |  |  | 0 (0) | 0 (0) | 0 (0) |  |
| Unknown | 6,672 (100) | 4,525 (100) | 2,147 (100) |  |  | 5,957 (100) | 4,588 (100) | 1,369 (100) |  |  | 485 (100) | 480 (100) | 5 (100) |  |
| **Atypical hyperplasia, n (%)** |  |  |  |  |  |  |  |  |  |  |  |  |  |  |
| No | 0 (0) | 0 (0) | 0 (0) |  |  | 0 (0) | 0 (0) | 0 (0) |  |  | 0 (0) | 0 (0) | 0 (0) |  |
| Yes | 0 (0) | 0 (0) | 0 (0) |  |  | 0 (0) | 0 (0) | 0 (0) |  |  | 0 (0) | 0 (0) | 0 (0) |  |
| Unknown | 6,672 (100) | 4,525 (100) | 2,147 (100) |  |  | 5,957 (100) | 4,588 (100) | 1,369 (100) |  |  | 485 (100) | 480 (100) | 5 (100) |  |
| **Median 5-year absolute risk by the Gail model (IQR)** | 0.594 (0.465 to 0.782) | 0.678 (0.549 to 0.828) | 0.465 (0.438 to 0.537) | <0.001 |  | 0.639 (0.465 to 0.836) | 0.759 (0.589 to 0.882) | 0.450 (0.424 to 0.477) | <0.001 |  | 0.750 (0.549 to 0.937) | 0.759 (0.565 to 0.938) | 0.462 (0.411 to 0.494) | 0.041 |
| **Protein truncating variants (9 Genes)** |  |  |  |  |  |  |  |  |  |  |  |  |  |  |
| No | 340 (5) | 340 (8) | 0 (0) |  |  | 744 (12) | 744 (16) | 0 (0) |  |  | 133 (27) | 133 (28) | 0 (0) |  |
| Yes | 6 (0) | 6 (0) | 0 (0) |  |  | 34 (1) | 34 (1) | 0 (0) |  |  | 2 (0) | 2 (0) | 0 (0) |  |
| Unknown | 6,326 (95) | 4,179 (92) | 2,147 (100) |  |  | 5,179 (87) | 3,810 (83) | 1,369 (100) |  |  | 350 (72) | 345 (72) | 5 (100) |  |
| **Polygenic risk score (PRS)** | 0.170 (-0.199 to 0.534) | 0.152 (-0.215 to 0.523) | 0.214 (-0.164 to 0.551) | 0.002 |  | 0.370 (0.000 to 0.752) | 0.357 (-0.018 to 0.737) | 0.399 (0.057 to 0.802) | <0.001 |  | 0.485 (0.116 to 0.841) | 0.484 (0.116 to 0.842) | 0.572 (0.421 to 0.731) | 0.89 |
| **Median 5-year absolute risk by PRS (IQR)** | 0.763 (0.515 to 1.122) | 0.748 (0.509 to 1.113) | 0.788 (0.529 to 1.143) | 0.004 |  | 0.931 (0.637 to 1.394) | 0.917 (0.623 to 1.374) | 0.974 (0.659 to 1.471) | <0.001 |  | 1.057 (0.720 to 1.558) | 1.057 (0.720 to 1.559) | 0.883 (0.876 to 1.476) | 0.835 |

#### **BCAC Funding and Acknowledgments**

**Funding**

This work was supported by Cancer Research UK grant: PPRPGM-Nov20\100002 and by core funding from the NIHR Cambridge Biomedical Research Centre (NIHR203312) [*]. *The views expressed are those of the author(s) and not necessarily those of the NIHR or the Department of Health and Social Care. Additional funding for BCAC is provided by the Confluence project which is funded with intramural funds from the National Cancer Institute Intramural Research Program, National Institutes of Health, the European Union's Horizon 2020 Research and Innovation Programme (grant numbers 634935 for BRIDGES), and the PERSPECTIVE I&I project, funded by the Government of Canada through Genome Canada and the Canadian Institutes of Health Research, the Ministère de l’Économie et de l'Innovation du Québec through Genome Québec, the Quebec Breast Cancer Foundation.

Genotyping of the OncoArray was funded by the NIH Grant U19 CA148065, and Cancer Research UK Grant C1287/A16563 and the PERSPECTIVE project supported by the Government of Canada through Genome Canada and the Canadian Institutes of Health Research (grant GPH-129344) and, the Ministère de l’Économie, Science et Innovation du Québec through Genome Québec and the PSRSIIRI-701 grant, and the Quebec Breast Cancer Foundation. Funding for iCOGS came from: the European Community's Seventh Framework Programme under grant agreement n° 223175 (HEALTH-F2-2009-223175) (COGS), Cancer Research UK (C1287/A10118, C1287/A10710, C12292/A11174, C1281/A12014, C5047/A8384, C5047/A15007, C5047/A10692, C8197/A16565), the National Institutes of Health (CA128978) and Post-Cancer GWAS initiative (1U19 CA148537, 1U19 CA148065 and 1U19 CA148112 - the GAME-ON initiative), the Department of Defence (W81XWH-10-1-0341), the Canadian Institutes of Health Research (CIHR) for the CIHR Team in Familial Risks of Breast Cancer, and Komen Foundation for the Cure, the Breast Cancer Research Foundation, and the Ovarian Cancer Research Fund.

The BRIDGES panel sequencing was supported by the European Union Horizon 2020 research and innovation program BRIDGES (grant number, 634935) and the Wellcome Trust (v203477/Z/16/Z).

The ABCFS was also supported by the National Health and Medical Research Council of Australia, the New South Wales Cancer Council, the Victorian Health Promotion Foundation (Australia) and the Victorian Breast Cancer Research Consortium. J.L.H. is a National Health and Medical Research Council (NHMRC) Senior Principal Research Fellow. M.C.S. is a NHMRC Senior Research Fellow. The ABCS study was supported by the Dutch Cancer Society [grants NKI 2007-3839; 2009 4363] and an institutional grant of the Dutch Cancer Society and of the Dutch Ministry of Health, Welfare and Sport. The Australian Breast Cancer Tissue Bank (ABCTB) was supported by the National Health and Medical Research Council of Australia, The Cancer Institute NSW and the National Breast Cancer Foundation. The ACP study is funded by the Breast Cancer Research Trust, UK. KM and AL are supported by the NIHR Manchester Biomedical Research Centre, the Allan Turing Institute under the EPSRC grant EP/N510129/1. The AHS study is supported by the intramural research program of the National Institutes of Health, the National Cancer Institute (grant number Z01-CP010119), and the National Institute of Environmental Health Sciences (grant number Z01-ES049030). The work of the BBCC was partly funded by ELAN-Fond of the University Hospital of Erlangen. The BBCS is funded by Cancer Research UK and Breast Cancer Now and acknowledges NHS funding to the NIHR Biomedical Research Centre, and the National Cancer Research Network (NCRN). The BCEES was funded by the National Health and Medical Research Council, Australia and the Cancer Council Western Australia (JS). For the BCFR-NY, BCFR-PA, BCFR-UT this work was supported by grant UM1 CA164920 from the National Cancer Institute. This work was supported by grant U01 CA164920 from the USA National Cancer Institute. The content of this manuscript does not necessarily reflect the views or policies of the National Cancer Institute or any of the collaborating centers in the Breast Cancer Family Registry (BCFR), nor does mention of trade names, commercial products, or organizations imply endorsement by the US Government or the BCFR. The BCINIS study is supported in part by the Breast Cancer Research Foundation (BCRF). The BREast Oncology GAlician Network (BREOGAN) is funded by Acción Estratégica de Salud del Instituto de Salud Carlos III FIS PI12/02125/Cofinanciado and FEDER PI17/00918/Cofinanciado FEDER; Acción Estratégica de Salud del Instituto de Salud Carlos III FIS Intrasalud (PI13/01136); Programa Grupos Emergentes, Cancer Genetics Unit, Instituto de Investigacion Biomedica Galicia Sur. Xerencia de Xestion Integrada de Vigo-SERGAS, Instituto de Salud Carlos III, Spain; Grant 10CSA012E, Consellería de Industria Programa Sectorial de Investigación Aplicada, PEME I + D e I + D Suma del Plan Gallego de Investigación, Desarrollo e Innovación Tecnológica de la Consellería de Industria de la Xunta de Galicia, Spain; Grant EC11-192. Fomento de la Investigación Clínica Independiente, Ministerio de Sanidad, Servicios Sociales e Igualdad, Spain; and Grant FEDER-Innterconecta. Ministerio de Economia y Competitividad, Xunta de Galicia, Spain. CBCS is funded by the Canadian Cancer Society (grant # 313404) and the Canadian Institutes of Health Research. CCGP is supported by funding from the University of Crete. The CECILE study was supported by Fondation de France, Institut National du Cancer (INCa), Ligue Nationale contre le Cancer, Agence Nationale de Sécurité Sanitaire, de l'Alimentation, de l'Environnement et du Travail (ANSES), Agence Nationale de la Recherche (ANR). The CGPS was supported by the Chief Physician Johan Boserup and Lise Boserup Fund, the Danish Medical Research Council, and Herlev and Gentofte Hospital. The American Cancer Society funds the creation, maintenance, and updating of the CPS-II cohort. The California Teachers Study (CTS) and the research reported in this publication were supported by the National Cancer Institute of the National Institutes of Health under award number U01-CA199277; P30-CA033572; P30-CA023100; UM1-CA164917; and R01-CA077398. The content is solely the responsibility of the authors and does not necessarily represent the official views of the National Cancer Institute or the National Institutes of Health. The collection of cancer incidence data used in the California Teachers Study was supported by the California Department of Public Health pursuant to California Health and Safety Code Section 103885; Centers for Disease Control and Prevention’s National Program of Cancer Registries, under cooperative agreement 5NU58DP006344; the National Cancer Institute’s Surveillance, Epidemiology and End Results Program under contract HHSN261201800032I awarded to the University of California, San Francisco, contract HHSN261201800015I awarded to the University of Southern California, and contract HHSN261201800009I awarded to the Public Health Institute. The opinions, findings, and conclusions expressed herein are those of the author(s) and do not necessarily reflect the official views of the State of California, Department of Public Health, the National Cancer Institute, the National Institutes of Health, the Centers for Disease Control and Prevention or their Contractors and Subcontractors, or the Regents of the University of California, or any of its programs. The University of Westminster curates the DietCompLyf database funded by Against Breast Cancer Registered Charity No. 1121258 and the NCRN. The coordination of EPIC is financially supported by the European Commission (DG-SANCO) and the International Agency for Research on Cancer. The national cohorts are supported by: Ligue Contre le Cancer, Institut Gustave Roussy, Mutuelle Générale de l’Education Nationale, Institut National de la Santé et de la Recherche Médicale (INSERM) (France); German Cancer Aid, German Cancer Research Center (DKFZ), Federal Ministry of Education and Research (BMBF) (Germany); the Hellenic Health Foundation, the Stavros Niarchos Foundation (Greece); Associazione Italiana per la Ricerca sul Cancro-AIRC-Italy and National Research Council (Italy); Dutch Ministry of Public Health, Welfare and Sports (VWS), Netherlands Cancer Registry (NKR), LK Research Funds, Dutch Prevention Funds, Dutch ZON (Zorg Onderzoek Nederland), World Cancer Research Fund (WCRF), Statistics Netherlands (The Netherlands); Health Research Fund (FIS), PI13/00061 to Granada, PI13/01162 to EPIC-Murcia, Regional Governments of Andalucía, Asturias, Basque Country, Murcia and Navarra, ISCIII RETIC (RD06/0020) (Spain); Cancer Research UK (14136 to EPIC-Norfolk; C570/A16491 and C8221/A19170 to EPIC-Oxford), Medical Research Council (1000143 to EPIC-Norfolk, MR/M012190/1 to EPIC-Oxford) (United Kingdom). The ESTHER study was supported by a grant from the Baden Württemberg Ministry of Science, Research and Arts. Additional cases were recruited in the context of the VERDI study, which was supported by a grant from the German Cancer Aid (Deutsche Krebshilfe). FHRISK and PROCAS are funded from NIHR grant PGfAR 0707-10031. DGE, AH and WGN are supported by the NIHR Manchester Biomedical Research Centre (IS-BRC-1215-20007). The GENICA was funded by the Federal Ministry of Education and Research (BMBF) Germany grants 01KW9975/5, 01KW9976/8, 01KW9977/0 and 01KW0114, the Robert Bosch Foundation, Stuttgart, Deutsches Krebsforschungszentrum (DKFZ), Heidelberg, the Institute for Prevention and Occupational Medicine of the German Social Accident Insurance, Institute of the Ruhr University Bochum (IPA), Bochum, as well as the Department of Internal Medicine, Johanniter GmbH Bonn, Johanniter Krankenhaus, Bonn, Germany. The GESBC was supported by the Deutsche Krebshilfe e. V. [70492] and the German Cancer Research Center (DKFZ). The HABCS study was supported by German Research Foundation (DFG Do761/15-1), the Claudia von Schilling Foundation for Breast Cancer Research, by the Lower Saxonian Cancer Society, and by the Rudolf Bartling Foundation. The HERPACC was supported by MEXT Kakenhi (No. 170150181 and 26253041) from the Ministry of Education, Science, Sports, Culture and Technology of Japan, by a Grant-in-Aid for the Third Term Comprehensive 10-Year Strategy for Cancer Control from Ministry Health, Labour and Welfare of Japan, by Health and Labour Sciences Research Grants for Research on Applying Health Technology from Ministry Health, Labour and Welfare of Japan, by National Cancer Center Research and Development Fund, and "Practical Research for Innovative Cancer Control (15ck0106177h0001 and 20ck0106553)" from Japan Agency for Medical Research and development, AMED, and Cancer Bio Bank Aichi. The HMBCS was supported by the German Research Foundation (DFG Do761/15-1), a grant from the Friends of Hannover Medical School, and by the Rudolf Bartling Foundation. ICICLE was supported by Breast Cancer Now, CRUK and Biomedical Research Centre at Guy’s and St Thomas’ NHS Foundation Trust and King’s College London. Financial support for KARBAC was provided through the regional agreement on medical training and clinical research (ALF) between Stockholm County Council and Karolinska Institutet, the Swedish Cancer Society, The Gustav V Jubilee foundation and Bert von Kantzows foundation. The KARMA study was supported by Märit and Hans Rausings Initiative Against Breast Cancer. The KBCP was financially supported by the special Government Funding (VTR) of Kuopio University Hospital grants, Cancer Fund of North Savo, the Finnish Cancer Organizations, and by the strategic funding of the University of Eastern Finland. kConFab is supported by a grant from the National Breast Cancer Foundation, and previously by the National Health and Medical Research Council (NHMRC), the Queensland Cancer Fund, the Cancer Councils of New South Wales, Victoria, Tasmania and South Australia, and the Cancer Foundation of Western Australia. Financial support for the AOCS was provided by the United States Army Medical Research and Materiel Command [DAMD17-01-1-0729], Cancer Council Victoria, Queensland Cancer Fund, Cancer Council New South Wales, Cancer Council South Australia, The Cancer Foundation of Western Australia, Cancer Council Tasmania and the National Health and Medical Research Council of Australia (NHMRC; 400413, 400281, 199600). G.C.T. and P.W. are supported by the NHMRC. RB was a Cancer Institute NSW Clinical Research Fellow. The KOHBRA study was supported by a grant from the National R&D Program for Cancer Control, Ministry of Health & Welfare, Republic of Korea (0720450; 1020350; 1420190) and the National R&D Program for Cancer Control through the National Cancer Center (NCC) funded by the Ministry of Health & Welfare, Republic of Korea (HA21C0140). LMBC is supported by the 'Stichting tegen Kanker'. DL is supported by the FWO. The MABCS study is funded by the Research Centre for Genetic Engineering and Biotechnology "Georgi D. Efremov", MASA. The MARIE study was supported by the Deutsche Krebshilfe e.V. [70-2892-BR I, 106332, 108253, 108419, 110826, 110828], the Hamburg Cancer Society, the German Cancer Research Center (DKFZ) and the Federal Ministry of Education and Research (BMBF) Germany [01KH0402]. The MCBCS was supported by the NIH grants R35CA253187, R01CA192393, R01CA116167, R01CA176785 a NIH Specialized Program of Research Excellence (SPORE) in Breast Cancer [P50CA116201], and the Breast Cancer Research Foundation. The Melbourne Collaborative Cohort Study (MCCS) cohort recruitment was funded by VicHealth and Cancer Council Victoria. The MCCS was further augmented by Australian National Health and Medical Research Council grants 209057, 396414 and 1074383 and by infrastructure provided by Cancer Council Victoria. Cases and their vital status were ascertained through the Victorian Cancer Registry and the Australian Institute of Health and Welfare, including the Australian Cancer Database. The MEC was supported by NIH grants CA63464, CA54281, CA098758, CA132839 and CA164973. The MISS study was supported by funding from ERC-2011-294576 Advanced grant, Swedish Cancer Society CAN 2018/675, Swedish Research Council, Local hospital funds, Berta Kamprad Foundation FBKS 2021-19, Gunnar Nilsson. The MMHS study was supported by NIH grants CA97396, CA128931, CA116201, CA140286 and CA177150. MSKCC is supported by grants from the Breast Cancer Research Foundation and Robert and Kate Niehaus Clinical Cancer Genetics Initiative. MYBRCA is funded by research grants from the Wellcome Trust (v203477/Z/16/Z), the Malaysian Ministry of Higher Education (UM.C/HlR/MOHE/06) and Cancer Research Malaysia. MYMAMMO is supported by research grants from Yayasan Sime Darby LPGA Tournament and Malaysian Ministry of Higher Education (RP046B-15HTM). The NBCS has received funding from the K.G. Jebsen Centre for Breast Cancer Research; the Research Council of Norway grant 193387/V50 (to A-L Børresen-Dale and V.N. Kristensen) and grant 193387/H10 (to A-L Børresen-Dale and V.N. Kristensen), South Eastern Norway Health Authority (grant 39346 to A-L Børresen-Dale) and the Norwegian Cancer Society (to A-L Børresen-Dale and V.N. Kristensen). The NBHS was supported by NIH grant R01CA100374. Biological sample preparation was conducted the Survey and Biospecimen Shared Resource, which is supported by P30 CA68485. The Carolina Breast Cancer Study (NCBCS) was funded by Komen Foundation, the National Cancer Institute (P50 CA058223, U54 CA156733, U01 CA179715), and the North Carolina University Cancer Research Fund. The NGOBCS was supported by the National Cancer Center Research and Development Fund (Japan). The NHS was supported by NIH grants P01 CA87969, UM1 CA186107, and U19 CA148065. The NHS2 was supported by NIH grants UM1 CA176726 and U19 CA148065. The ORIGO study was supported by the Dutch Cancer Society (RUL 1997-1505) and the Biobanking and Biomolecular Resources Research Infrastructure (BBMRI-NL CP16). The PBCS was funded by Intramural Research Funds of the National Cancer Institute, Department of Health and Human Services, USA. Genotyping for PLCO was supported by the Intramural Research Program of the National Institutes of Health, NCI, Division of Cancer Epidemiology and Genetics. The PLCO is supported by the Intramural Research Program of the Division of Cancer Epidemiology and Genetics and supported by contracts from the Division of Cancer Prevention, National Cancer Institute, National Institutes of Health. The SASBAC study was supported by funding from the Agency for Science, Technology and Research of Singapore (A*STAR), the US National Institute of Health (NIH) and the Susan G. Komen Breast Cancer Foundation. The SBCGS was supported primarily by NIH grants R01CA64277, R01CA148667, UMCA182910, and R37CA70867. Biological sample preparation was conducted the Survey and Biospecimen Shared Resource, which is supported by P30 CA68485. The scientific development and funding of this project were, in part, supported by the Genetic Associations and Mechanisms in Oncology (GAME-ON) Network U19 CA148065. The SBCS was supported by Sheffield Experimental Cancer Medicine Centre and Breast Cancer Now Tissue Bank. SEARCH was funded by Cancer Research UK [C490/A10124, C490/A16561] and supported by the UK National Institute for Health Research Biomedical Research Centre at the University of Cambridge. SEBCS was supported by the BRL (Basic Research Laboratory) program through the National Research Foundation of Korea funded by the Ministry of Education, Science and Technology (2012-0000347). SGBCC is funded by the National Research Foundation Singapore, NUS start-up Grant, National University Cancer Institute Singapore (NCIS) Centre Grant, Breast Cancer Prevention Programme, Asian Breast Cancer Research Fund and the NMRC Clinician Scientist Award (SI Category). Population-based controls were from the Multi-Ethnic Cohort (MEC) funded by grants from the Ministry of Health, Singapore, National University of Singapore and National University Health System, Singapore. The Sister Study (SISTER) is supported by the Intramural Research Program of the NIH, National Institute of Environmental Health Sciences (Z01-ES044005 and Z01-ES049033). The Two Sister Study (2SISTER) was supported by the Intramural Research Program of the NIH, National Institute of Environmental Health Sciences (Z01-ES044005 and Z01-ES102245), and, also by a grant from Susan G. Komen for the Cure, grant FAS0703856. SKKDKFZS is supported by the DKFZ. SMC is part of the Swedish Infrastructure for Medical Population-based Life-course and Environmental Research (SIMPLER), which receives funding from the Swedish Research Council (grants VR 2017-00644, VR 2021-00160). The study is supported by the Swedish Cancer Foundation grant CF 20 0864. The TWBCS is supported by the Taiwan Biobank project of the Institute of Biomedical Sciences, Academia Sinica, Taiwan. UBCS was supported by funding from National Cancer Institute (NCI) grant R01 CA163353 (to N.J. Camp) and the Women’s Cancer Center at the Huntsman Cancer Institute (HCI). Data collection for UBCS was supported by the Utah Population Database (UPDB) and Utah Cancer Registry (UCR). The UPDB is supported by HCI, the University of Utah, and NCI grant P30 CA2014. The UCR is additionally funded by the NCI's SEER Program, HHSN261201800016I, and the US Center for Disease Control and Prevention's National Program of Cancer Registries (NU58DP007131). The UCIBCS component of this research was supported by the NIH [CA58860, CA92044] and the Lon V Smith Foundation [LVS39420]. The UKBGS is funded by Breast Cancer Now and the Institute of Cancer Research (ICR), London. ICR acknowledges NHS funding to the NIHR Biomedical Research Centre. The USRT Study was funded by Intramural Research Programof the National Cancer Institute, Department of Health and Human Services, USA. This work was also funded by NCI U19 CA148065-01.

**Acknowledgements**

We thank all the individuals who took part in these studies and all the researchers, clinicians, technicians and administrative staff who have enabled this work to be carried out. ABCFS thank Maggie Angelakos, Judi Maskiell, Gillian Dite. ABCS thanks the Blood bank Sanquin, The Netherlands. ABCTB Investigators: Christine Clarke, Deborah Marsh, Rodney Scott, Robert Baxter, Desmond Yip, Jane Carpenter, Alison Davis, Nirmala Pathmanathan, Peter Simpson, J. Dinny Graham, Mythily Sachchithananthan. Samples are made available to researchers on a non-exclusive basis. The ACP study wishes to thank the participants in the Thai Breast Cancer study. Special thanks also go to the Thai Ministry of Public Health (MOPH), doctors and nurses who helped with the data collection process. Finally, the study would like to thank Dr Prat Boonyawongviroj, the former Permanent Secretary of MOPH and Dr Pornthep Siriwanarungsan, the former Department Director-General of Disease Control who have supported the study throughout. BBCS thanks Eileen Williams, Elaine Ryder-Mills, Kara Sargus. BCEES thanks Allyson Thomson, Christobel Saunders, Jennifer Girschik, Jane Heyworth and Terry Boyle. The BCINIS study would not have been possible without the major contribution of Ms. H. Rennert, and the contributions of Dr. M. Pinchev, Dr. O. Barnet, Dr. N. Gronich, Dr. K. Landsman, Dr. A. Flugelman, Dr. W. Saliba, Dr. E. Liani, Dr. I. Cohen, Dr. S. Kalet, Dr. V. Friedman of the NICCC in Haifa, and all the contributing family medicine, surgery, pathology and oncology teams in all medical institutes in Northern Israel. The BREOGAN study would not have been possible without the contributions of the following: Manuela Gago-Dominguez, Jose Esteban Castelao, Angel Carracedo, Victor Muñoz Garzón, Alejandro Novo Domínguez, Maria Elena Martinez, Sara Miranda Ponte, Carmen Redondo Marey, Maite Peña Fernández, Manuel Enguix Castelo, Maria Torres, Manuel Calaza (BREOGAN), José Antúnez, Máximo Fraga and the staff of the Department of Pathology and Biobank of the University Hospital Complex of Santiago-CHUS, Instituto de Investigación Sanitaria de Santiago, IDIS, Xerencia de Xestion Integrada de Santiago-SERGAS; Joaquín González-Carreró and the staff of the Department of Pathology and Biobank of University Hospital Complex of Vigo, Instituto de Investigacion Biomedica Galicia Sur, SERGAS, Vigo, Spain. CGPS thanks staff and participants of the Copenhagen General Population Study. For the excellent technical assistance: Dorthe Uldall Andersen, Maria Birna Arnadottir, Anne Bank, Dorthe Kjeldgård Hansen. The Danish Cancer Biobank is acknowledged for providing infrastructure for the collection of blood samples for the cases. The Danish Breast Cancer Cooperative Group (DBCG) are acknowledged for their provision of clinical case data. CNIO-BCS thanks Guillermo Pita, Charo Alonso, Nuria Álvarez, Pilar Zamora, Primitiva Menendez, the Human Genotyping-CEGEN Unit (CNIO). COLBCCC thanks all patients, the physicians Justo G. Olaya, Mauricio Tawil, Lilian Torregrosa, Elias Quintero, Sebastian Quintero, Claudia Ramírez, José J. Caicedo, and Jose F. Robledo, and the technician Michael Gilbert for their contributions and commitment to this study. Investigators from the CPS-II cohort thank the participants and Study Management Group for their invaluable contributions to this research. They also acknowledge the contribution to this study from central cancer registries supported through the Centers for Disease Control and Prevention National Program of Cancer Registries, as well as cancer registries supported by the National Cancer Institute Surveillance Epidemiology and End Results program. The authors would like to thank the California Teachers Study Steering Committee that is responsible for the formation and maintenance of the Study within which this research was conducted. A full list of California Teachers Study (CTS) team members is available at https://www.calteachersstudy.org/team. DIETCOMPLYF thanks the patients, nurses and clinical staff involved in the study. The DietCompLyf study was funded by the charity Against Breast Cancer (Registered Charity Number 1121258) and the NCRN. We thank the participants and the investigators of EPIC (European Prospective Investigation into Cancer and Nutrition). ESTHER thanks Hartwig Ziegler, Sonja Wolf, Volker Hermann, Christa Stegmaier, Katja Butterbach. FHRISK and PROCAS thank NIHR for funding. The GENICA Network: Dr. Margarete Fischer-Bosch-Institute of Clinical Pharmacology, Stuttgart, and University of Tübingen, Germany [Hiltrud Brauch, RH, Wing-Yee Lo], Department of Internal Medicine, Johanniter GmbH Bonn, Johanniter Krankenhaus, Bonn, Germany [YDK, Christian Baisch], Institute of Pathology, University of Bonn, Germany [Hans-Peter Fischer], Molecular Genetics of Breast Cancer, Deutsches Krebsforschungszentrum (DKFZ), Heidelberg, Germany [UH], Institute for Prevention and Occupational Medicine of the German Social Accident Insurance, Institute of the Ruhr University Bochum (IPA), Bochum, Germany [Thomas Brüning, Beate Pesch, Sylvia Rabstein, Anne Lotz]; and Institute of Occupational Medicine and Maritime Medicine, University Medical Center Hamburg-Eppendorf, Germany [Volker Harth]. HKBCS thanks Hong Kong Sanatorium and Hospital, Dr Ellen Li Charitable Foundation, The Kerry Group Kuok Foundation for support. HMBCS thanks Peter Hillemanns, Hans Christiansen and Johann H. Karstens. HUBCS thanks Darya Prokofyeva and Shamil Gantsev. ICICLE thanks Kelly Kohut, Michele Caneppele, Maria Troy. KARMA and SASBAC thank the Swedish Medical Research Counsel. KBCP thanks Eija Myöhänen. kConFab/AOCS wish to thank Heather Thorne, Eveline Niedermayr, all the kConFab research nurses and staff, the heads and staff of the Family Cancer Clinics, and the Clinical Follow Up Study (which has received funding from the NHMRC, the National Breast Cancer Foundation, Cancer Australia, and the National Institute of Health (USA)) for their contributions to this resource, and the many families who contribute to kConFab. We thank all investigators of the KOHBRA (Korean Hereditary Breast Cancer) Study. LMBC thanks Gilian Peuteman, Thomas Van Brussel, EvyVanderheyden and Kathleen Corthouts. MABCS thanks Milena Jakimovska (RCGEB “Georgi D. Efremov”), Snezhana Smichkoska, Emilija Lazarova, Marina Iljoska (University Clinic of Radiotherapy and Oncology), Katerina Kubelka-Sabit, Dzengis Jasar, Mitko Karadjozov (Adzibadem-Sistina Hospital), Andrej Arsovski and Liljana Stojanovska (Re-Medika Hospital) for their contributions and commitment to this study. MARIE thanks Petra Seibold, Ursula Eilber and Muhabbet Celik. MBCSG (Milan Breast Cancer Study Group): Paolo Radice, Paolo Peterlongo, Siranoush Manoukian, Bernard Peissel, Jacopo Azzollini, Erica Rosina, Daniela Zaffaroni, Bernardo Bonanni, Irene Feroce, Mariarosaria Calvello, Aliana Guerrieri Gonzaga, Monica Marabelli, Davide Bondavalli and the personnel of the Cogentech Cancer Genetic Test Laboratory. The MCCSs was made possible by the contribution of many people, including the original investigators, the teams that recruited the participants and continue working on follow-up, and the many thousands of Melbourne residents who continue to participate in the study. The MISS study group acknowledges the former Principal Investigator, Professor Håkan Olsson. We thank the coordinators, the research staff and especially the MMHS participants for their continued collaboration on research studies in breast cancer. MSKCC thanks Marina Corines, Lauren Jacobs. NBHS and SBCGS thank study participants and research staff for their contributions and commitment to the studies. MyBrCa Investigators: Prof Kartini Rahmat, Prof See Mee Hoong, Associate Prof Suniza Jamaris, Dr. Tania Islam, Dr. Hamizah Saat, Dr. Jeannie Wong Hsiu Ding, Dr Tai Mei Chee, Dr Shivaani Mariapun, Ms Yoon Sook-Yee. For NHS and NHS2 the study protocol was approved by the institutional review boards of the Brigham and Women’s Hospital and Harvard T.H. Chan School of Public Health, and those of participating registries as required. We would like to thank the participants and staff of the NHS and NHS2 for their valuable contributions as well as the following state cancer registries for their help: AL, AZ, AR, CA, CO, CT, DE, FL, GA, ID, IL, IN, IA, KY, LA, ME, MD, MA, MI, NE, NH, NJ, NY, NC, ND, OH, OK, OR, PA, RI, SC, TN, TX, VA, WA, WY. The authors assume full responsibility for analyses and interpretation of these data. PREFACE thanks Sonja Oeser and Silke Landrith. The RBCS thanks Jannet Blom, Saskia Pelders, Wendy J.C. Prager – van der Smissen, and the Erasmus MC Family Cancer Clinic. SBCS thanks Sue Higham, Helen Cramp, Dan Connley, Ian Brock, Sabapathy Balasubramanian and Malcolm W.R. Reed. We thank the SEARCH and EPIC teams. SGBCC thanks the participants and all research coordinators for their excellent help with recruitment, data and sample collection. SGBCC Investigators: Benita Kiat-Tee Tan, Veronique Kiak Mien Tan, Geok Hoon Lim^,^ Ern Yu Tan, Su-Ming Tan. SKKDKFZS thanks all study participants, clinicians, family doctors, researchers and technicians for their contributions and commitment to this study. We thank the SUCCESS Study teams in Munich, Duessldorf, Erlangen and Ulm. UBCS thanks all study participants as well as the ascertainment, laboratory, analytics and informatics teams at Huntsman Cancer Institute and Intermountain Healthcare for their important contributions to this study. UCIBCS thanks Irene Masunaka. UKBGS thanks Breast Cancer Now and the Institute of Cancer Research for support and funding of the Generations Study, and the study participants, study staff, and the doctors, nurses and other health care providers and health information sources who have contributed to the study. We acknowledge NHS funding to the Royal Marsden/ICR NIHR Biomedical Research Centre.
